# Seroprevalence of COVID-19 in Palestine in 2020

**DOI:** 10.1101/2021.10.04.21263131

**Authors:** I Rayan, SE Qaddomi, O Najjar, S Abbas, K Mousa, L Iraqi, E Abdelkreem Aly, K Abu Khader, A Barakat, R Salman

## Abstract

COVID-19 affected different countries in different ways. Palestine had recorded over 140,000 cases by the end of 2020. The WHO/PNIPH, WHO/EMRO, and the Palestinian MoH carried out a serological survey in Palestine in order to estimate the actual number of COVID-19 infections up to the end of December 2020. A sample stratified by region, district, residence area (urban, rural, and refugee camp), and accounting for gender, was taken from Gaza and the West Bank. Data from participants were also collected, including demographic, socio-economic, and health conditions. The results show that 39% of the Palestinian population (38% of the West Bank and 40% of Gaza) had been infected with COVID-19 by the end of December, almost 10 times the number detected by targeted Rt-PCR testing. Several factors were calculated to be significant such as diabetes, smoking, gender, age, and residence.

**Summary of findings:** The following table is a summary of all findings presented in this report. The P values in green are below 0.05, which makes the result statistically significant; red is not statistically significant. In binary comparisons (when comparing two numbers), the odds were calculated, meaning how much more likely the presence of seropositivity is if the condition is satisfied. For example, those who were previously diagnosed as COVID-19 positive using Rt PCR were 2.5 times as likely to be seropositive than those who were not diagnosed.

| Correlation | P Value | Odds |
| --- | --- | --- |
| Distribution of Cases by Age Group (West Bank) | 0.00001 |  |
| Distribution of Cases by Age Group (Gaza Strip) | 0.00001 |  |
| Distribution of Cases by Gender (West Bank) | 0.00001 |  |
| Distribution of Cases by Gender (Gaza Strip) | 0.00001 |  |
| **Demographic Factors** |  |  |
| Residence Area type | 0.00001 |  |
| Marital Status | 0.0167 |  |
| Distribution of Cases by Region | 0.00001 |  |
| Distribution of Cases by District | 0.00001 |  |
| Rooms in Household (more than three) | 0.0141 | 1.11 <u>less</u> likely to be positive |
| Females in Household (more than three) | 0.0001 | 1.21 more likely to be positive |
| Males in Household (more than three) | 0.9755 | No Effect |
| Job Sector | 0.0001 |  |
| Traveling to Israel Often | 0.0022 | 1.33 <u>less</u> likely to be positive |
| Employment Status (Males) | 0.9159 | No Effect |
| Employment Status (Females) | 0.9302 | No Effect |
| **Underlying Conditions and Risk Factors** |  |  |
| Underlying Conditions | 0.3579 | 1.05 <u>less</u> likely to be positive |
| Diabetes | 0.0001 | 1.18 more likely to be positive |
| Hypertension | 0.5359 | 1.03 more likely to be positive |
| Cancer | 0.1832 | 1.31 more likely to be positive |
| Pregnancy | 0.4833 | 1.18 more likely to be positive |
| Smoking | 0.0001 | 1.63 <u>less</u> likely to be positive |
| **Symptoms and Detection** |  |  |
| Previously PCR Positive | 0.00001 | 2.50 more likely to be positive |
| Symptomatic | 0.00001 | 1.70 more likely to be positive |
| Missing Work Due to Symptoms | 0.0001 | 1.38 more likely to be positive |
| Requiring Medical Attention for Symptoms | 0.0001 | 1.51 more likely to be positive |
| Requiring Hospitalization for Symptoms | 0.0221 | 1.25 more likely to be positive |
| Self-Reported COVID Contact | 0.0022 | 1.79 more likely to be positive |
| **Hospitalization Indicators** |  |  |
| Hospitalization by Age | 0.0001 |  |
| Hospitalization of Diabetics | 0.0001 | 3.55 more likely to be positive |

## Introduction

### Geographical Background

The physical separation of the West Bank and Gaza Strip limits travel and connection between them, thereby creating two distinct epidemiological realities. The West Bank has a large number of Palestinians who enter and exit areas controlled by Israel, while Gaza is generally isolated with barely any travel into areas controlled by Israel. Gaza has a much denser population than the West Bank of 5590.4 persons per km^2^ as opposed to 540.7 persons per km^2^ in the West Bank.

**Figure 1.1.**
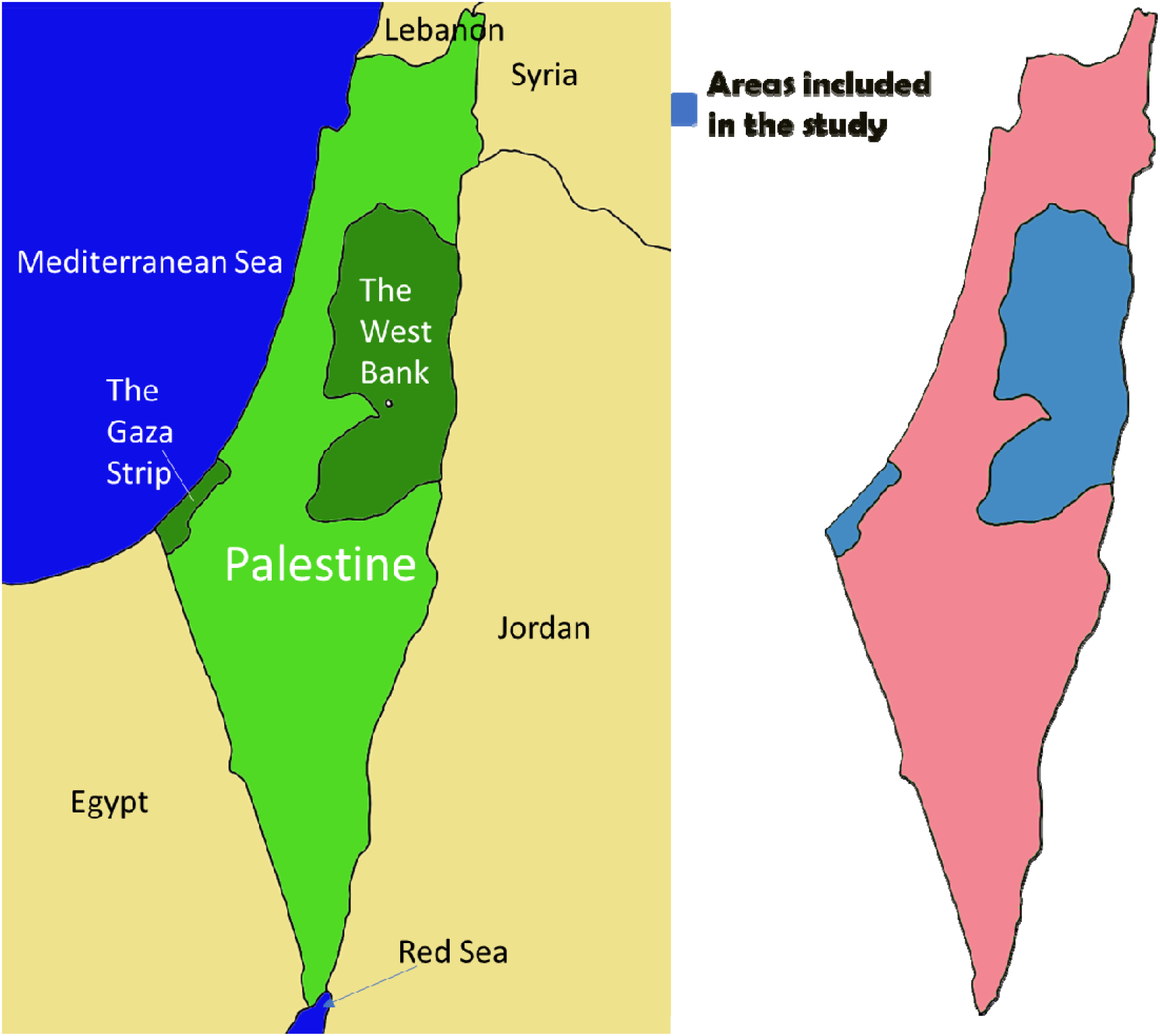
Regional map of Palestine (left) and Palestine showing the oPt (right).

Officially, the West Bank includes disputed East Jerusalem, which is mainly populated by Palestinians. However, most of the Palestinians in East Jerusalem are not technically Palestinian citizens, nor do they fall under the authority of the Palestinian state and are not affected by the public health measures undertaken by the Palestinian Ministry of Health (MoH). Therefore, for the purposes of this report, Jerusalem will refer to the localities in Jerusalem that actually do fall under the Palestinian Authority (outside the Wall).

### Demographic Background

According to the Palestinian Central Bureau of Statistics (PCBS), Palestine has a population of 5,101,152 (mid- year 2020), with 3,053,1 83 in the West Bank and 2,047,969 in the Gaza Strip. Palestine has a very a young population with 69% of the population under the age of 24. The current growth rate in Palestine is estimated to be 2.23% annually. This is a significant factor in any epidemic, and especially COVID-19 which affects age groups in different ways.

**Figure 1.2.**
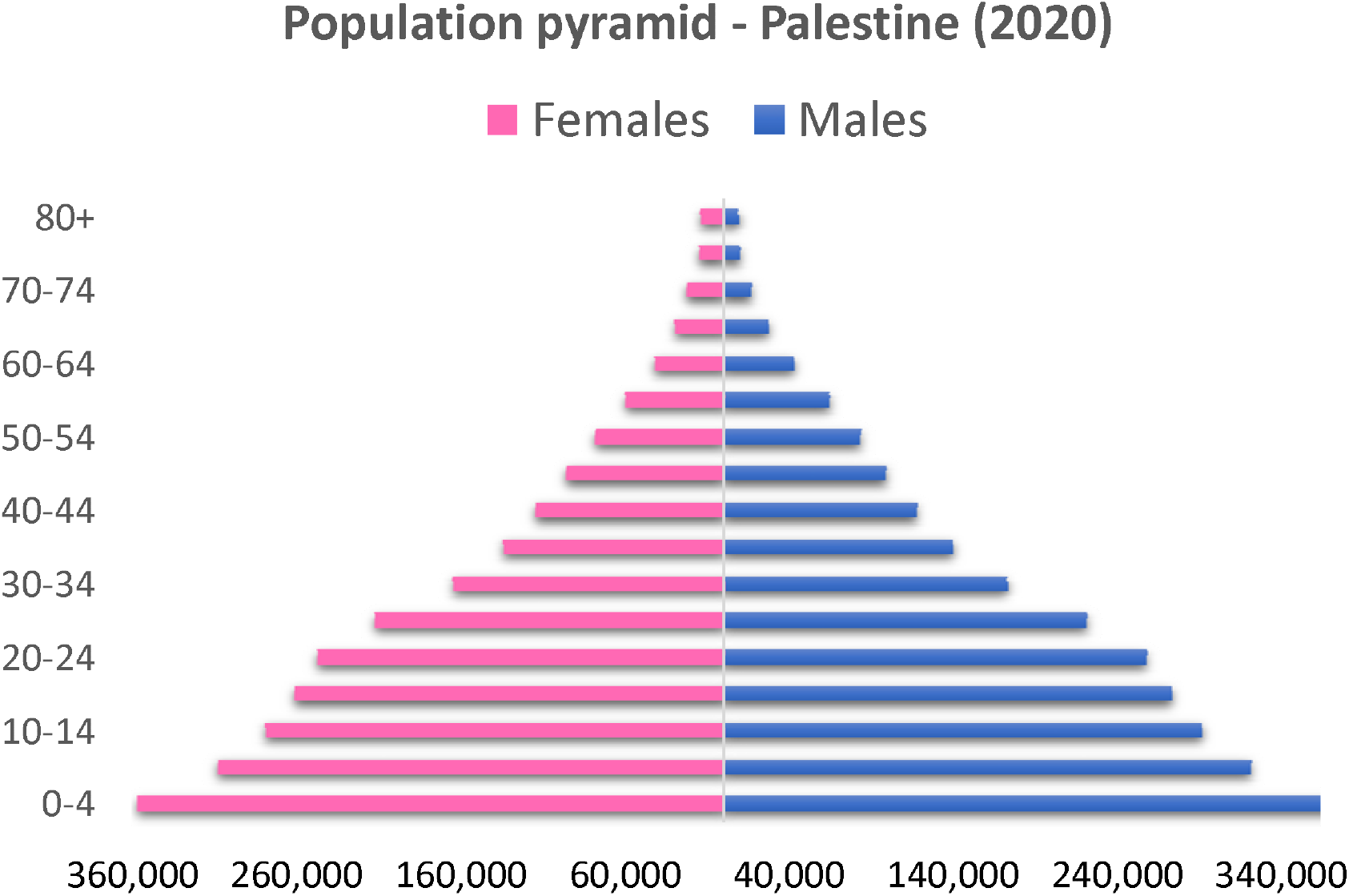
Palestine Population Pyramid (2020)

The West Bank pyramid is slightly more constricted than that of Gaza and shows that 67% of the population are below 30 years of age, while Gaza’s more expansive pyramid shows that 72% of the population are under the age of 30. In other words, Gaza’s population is much younger than that of the West Bank.

**Figure 1.3.**
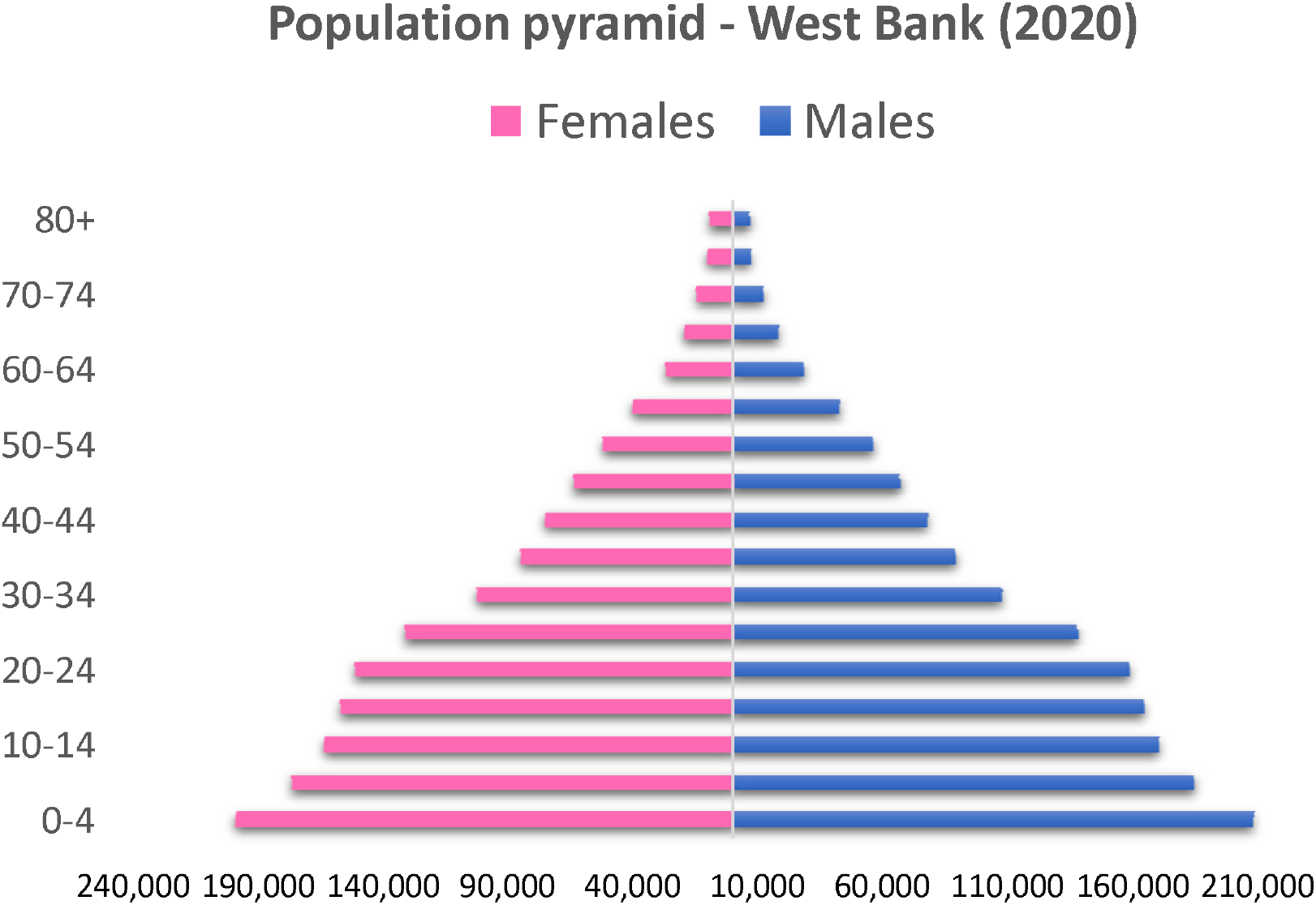
West Bank Population Pyramid (2020)

**Figure 1.4.**
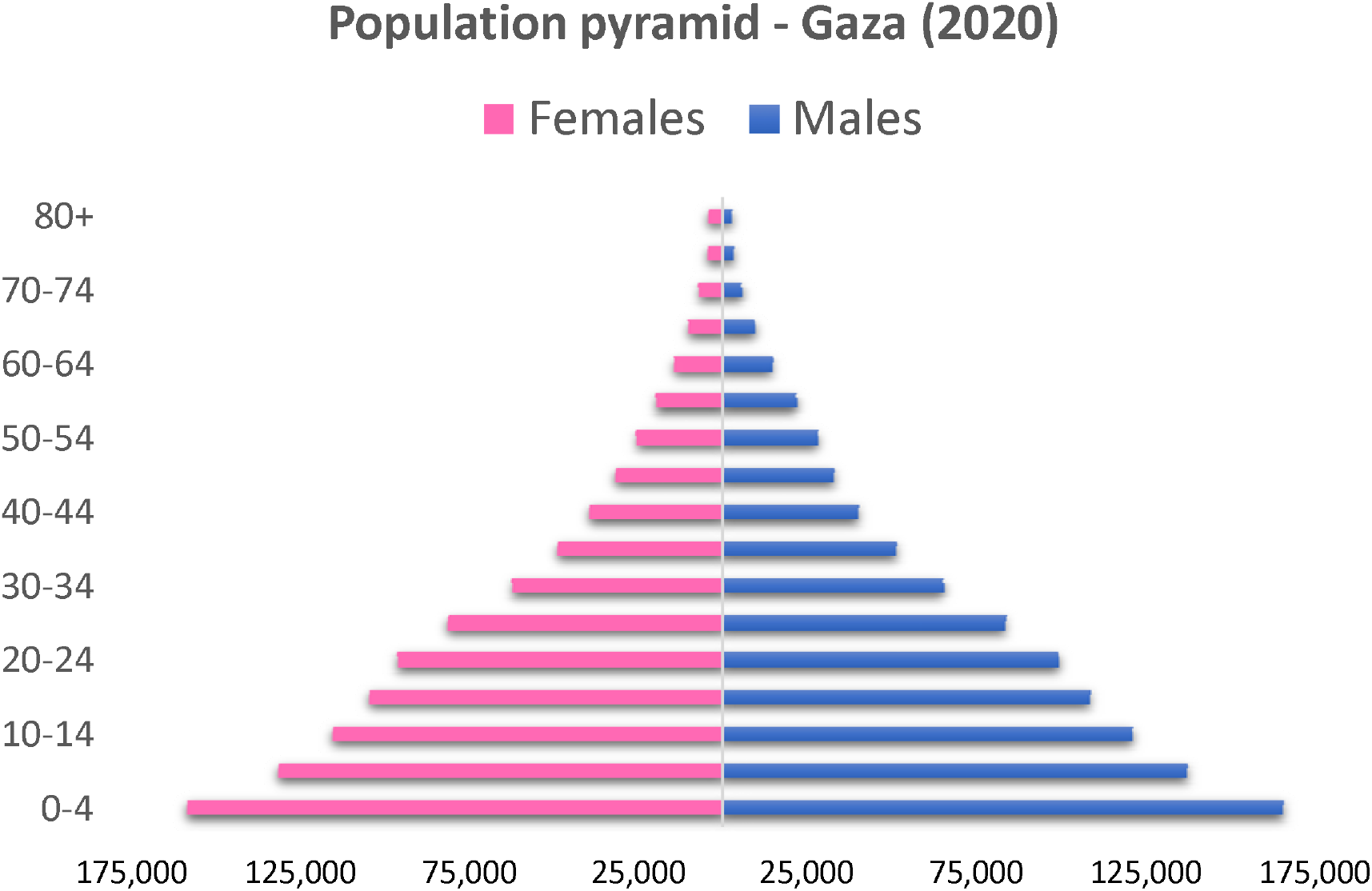
Gaza Population Pyramid (2020)

### COVID-19 on a Global Scale

COVID-19 began its spread from Wuhan, China, in December 2019. Wuhan was closed down in January 2020 in an attempt to limit the spread of the virus, an attempt which may have been too late as cases began to appear in many other countries soon after. Individual countries adopted measures of varying severity after the World Health Organization declared COVID a global pandemic in March 2020. The announcement prompted most nations to close their borders and forbid air travel.

Global figures indicate that by May 21, 2021, more than 168 million cases of COVID-19 infections had been reported with more than 3.49 million fatalities. Nations adopted various strategies to limit the spread of the virus, including monitoring new cases and isolating them, increasing the capacity and efficiency of their healthcare systems, redistribution of their health work force, and replenishing vital supplies.

### COVID-19 in Palestine

COVID-19 put an additional strain on the Palestinian healthcare system, which has historically suffered from multiple issues, exacerbated by the Israeli occupation, a lack of funding, and the recent decline of international support.

The COVID-19 epidemic has had an impact on governance, public health perceptions, and has changed the world in many ways. Palestine is no exception to this rule as it has been and continues to experience waves of infection.

To date (May 21, 2021), the Ministry of Health has detected a total of 307,536 cases in Palestine: 198,925 in the West Bank and 108,611 in the Gaza Strip.

**Figure 1.5.**
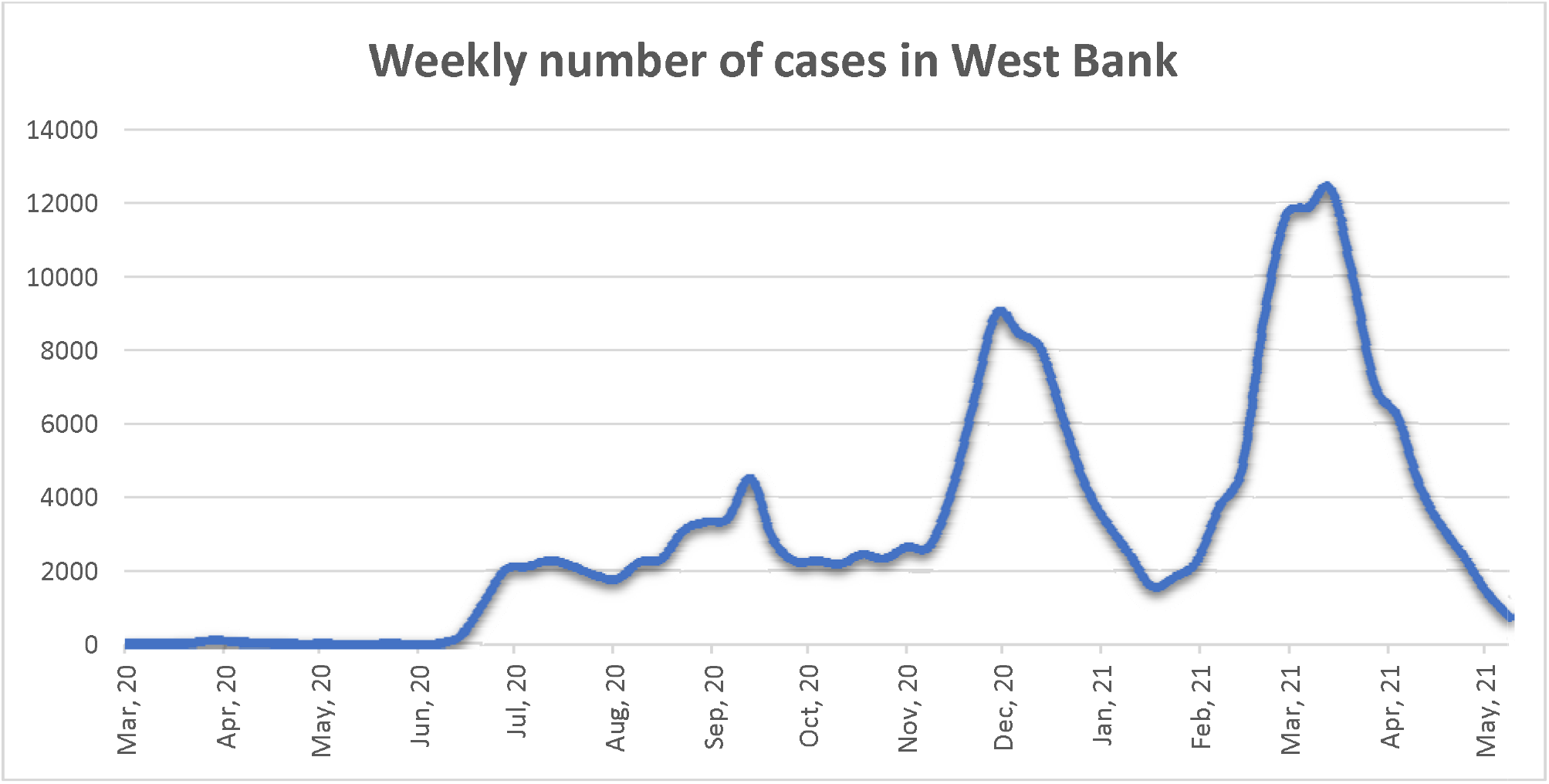
Weekly Number of Cases in the West Bank

**Figure 1.6.**
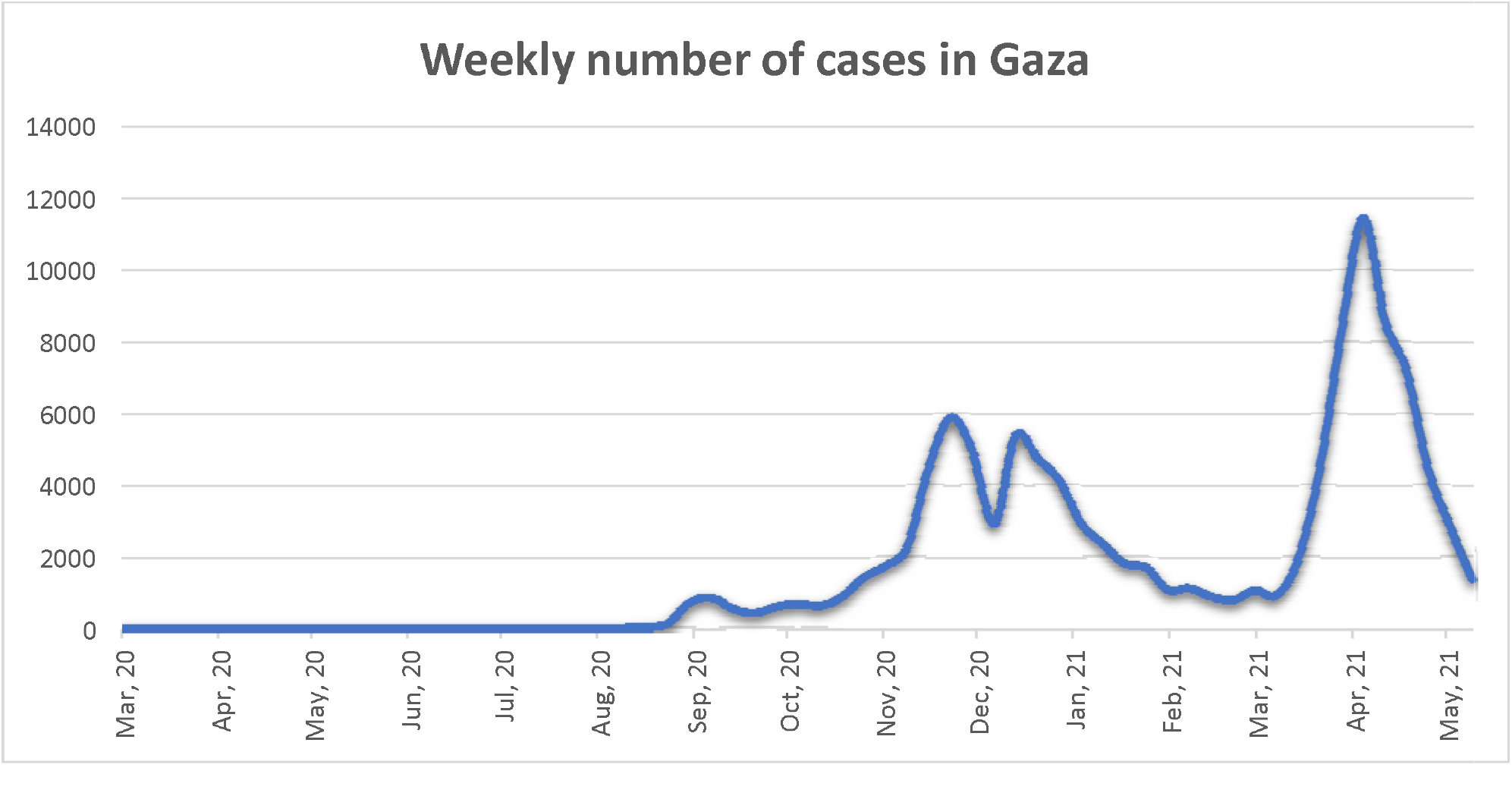
Weekly Number of Cases in Gaza

Since the first cases were detected in Palestine in March 2020, testing strategy in the West Bank and Gaza has relied on testing suspected cases (either those who came into contact with positive individuals or those showing symptoms). This was reflected in the overall positivity rate of 14.3% for the period. The number of undiagnosed or unreported cases was, therefore, unknowable.

### Study Objective

The World Health Organization East Mediterranean Regional Office (WHO/EMRO) spearheaded multiple serological surveys in the region as part of a larger serological project that included 54 countries worldwide. The WHO/PNIPH cooperated with the Palestinian Ministry of Health (MoH) to run a serological scan of Palestine.

The survey was initiated in the West Bank and Gaza to estimate a more accurate number of cases, including cases undetected by other methods, through random serological testing that can detect immune cells that result from infection; these immune cells can remain in the body for up to eight months. This type of study promised to shed light on the true epidemiological situation in Palestine.

## Methodology

### Sampling

The sample for the project was selected with an expected prevalence of 50%, a margin of error of 5%, a confidence interval of 95%, and a design effect of one.

The West Bank and Gaza were divided into three regions: north, middle, and south, and each region was stratified into three types: urban, rural, and camp. In Gaza the regions were divided into two types: urban and camp. Gaza has virtually no rural areas due to its high population density. Thus, there were six strata for the West Bank and five for the Gaza Strip.

The total sample size for Palestine was 7260.

|  |  |
| --- | --- |
| **West Bank (6 strata)** | **total sample size= 4356** |
| **For Gaza Strip (5 strata)** | **total sample size = 2904** |
| **Total number of sample size** | **Palestine (7260)** |

A total of 6063 random households were selected in the West Bank and Gaza (3638 in the West Bank and 2425 in the Gaza Strip) with assistance from the PCBS. Two blood samples (one from a male and one from a female) were taken from each household and sent to a MoH lab for analysis. The blood samples were accompanied by comprehensive forms containing as much information as possible regarding the individuals who supplied the samples for analysis.

### 13 in WB and 5 in GS (3 people 2 collectors and one statistician, questionnaire, add questionnaire to annex, random household)

#### Ethical consideration and consent form

##### This project was approved by the WHO/EMRO ethics board

###### Inclusion criteria

All individuals over the age of 10 years irrespective of known prior COVID-19 infection who resided in the area under study during the period of SARS-CoV-2 transmission and who can give consent, or in the case of minor children, whose legal guardian can give consent for them.

###### Exclusion criteria

All individuals who refuse or are unable to give informed consent, or who present contraindication to venipuncture. Prisoners were excluded due to logistical difficulties. Individuals with conditions that make sampling them potentially harmful and persons currently infected with COVID-19.

The testing kit used was a Roche Elecsys Anti-SARS-CoV-2 S Immunoassay used for the in-vitro quantitative determination of antibodies (including IgG) to the Severe Acute Respiratory Syndrome Coronavirus 2 (SARS⍰CoV⍰2) spike (S) protein receptor binding domain (RBD) in human serum and plasma. The cutoff point was 0.80 U/ml. The test specificity was 99.98 % and test sensitivity was 98.8 %. Samples were stable for 3 days at 15⍰25 °C or for 14 days at 2⍰8 °C.

## Results

### Demographic Factors Results

The survey showed that 38% of the West Bank and 40% of Gaza were seropositive, meaning that these individuals had been infected since March 2020. These rates were almost 10 times higher than the number of cases detected by the national surveillance system for COVID-19. The distribution of the prevalence was far from homogenous, with some districts showing 65% while others showed 15%.

The following are the results from the West Bank and Gaza divided by gender and age.

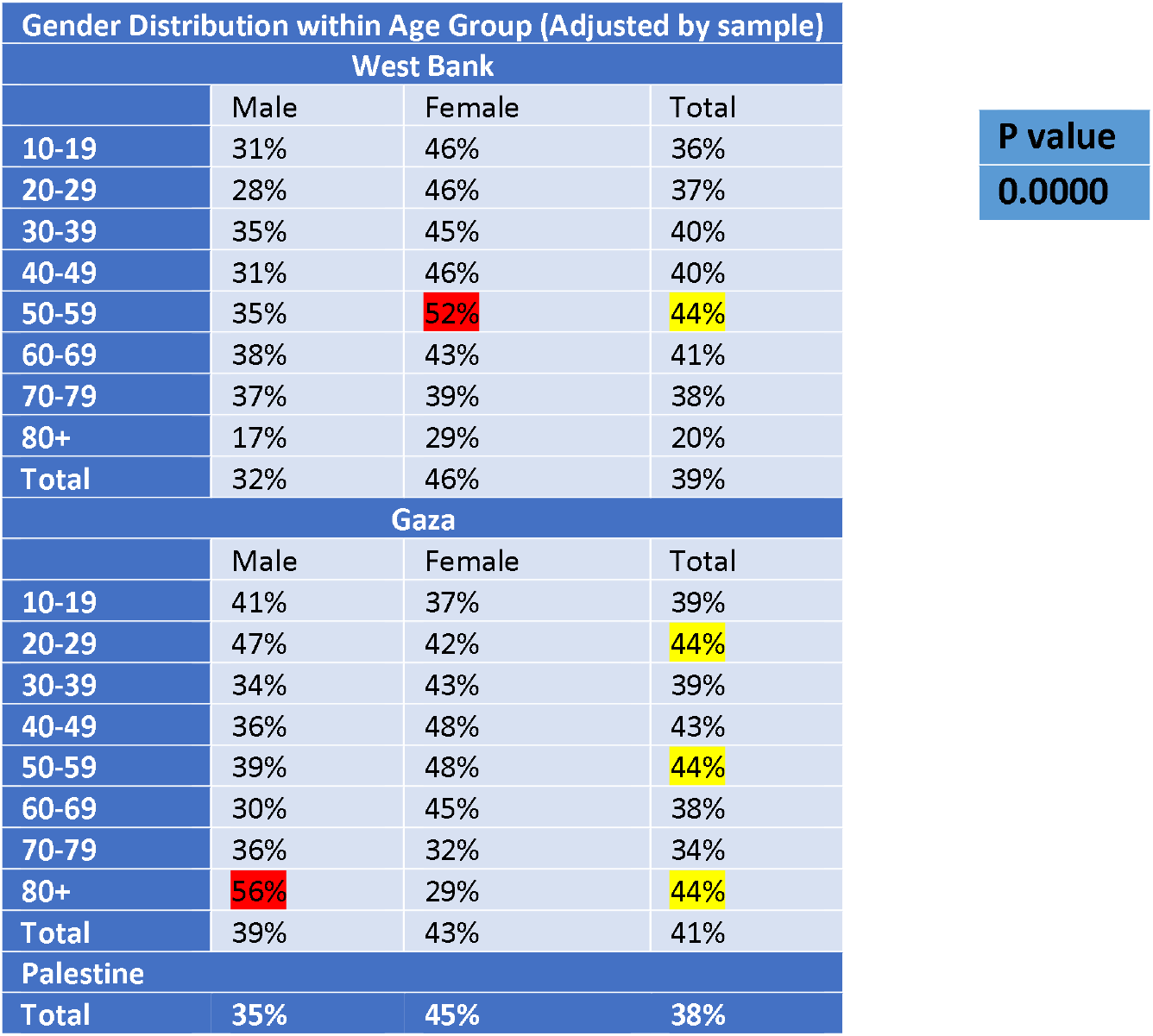

| Gender Distribution within Age Group (Adjusted by sample) |  |  |  |
| --- | --- | --- | --- |
| West Bank |  |  |  |
|  | Male | Female | Total |
| 10-19 | 31% | 46% | 36% |
| 20-29 | 28% | 46% | 37% |
| 30-39 | 35% | 45% | 40% |
| 40-49 | 31% | 46% | 40% |
| 50-59 | 35% | 52% | 44% |
| 60-69 | 38% | 43% | 41% |
| 70-79 | 37% | 39% | 38% |
| 80+ | 17% | 29% | 20% |
| Total | 32% | 46% | 39% |
| Gaza |  |  |  |
|  | Male | Female | Total |
| 10-19 | 41% | 37% | 39% |
| 20-29 | 47% | 42% | 44% |
| 30-39 | 34% | 43% | 39% |
| 40-49 | 36% | 48% | 43% |
| 50-59 | 39% | 48% | 44% |
| 60-69 | 30% | 45% | 38% |
| 70-79 | 36% | 32% | 34% |
| 80+ | 56% | 29% | 44% |
| Total | 39% | 43% | 41% |
| Palestine |  |  |  |
| Total | 35% | 45% | 38% |

P value

(Table) Distribution of seroprevalence in the West Bank and Gaza by age and gender. The most prevalent groups are highlighted.

In the West Bank, females had higher seroprevalence across all age groups. The 50–59-year age group had the highest prevalence (highlighted in yellow).

In Gaza, females also had higher seroprevalence, albeit less than in the West Bank. The 50-59 age group also had the highest prevalence, along with the 20-29 and 80+ age groups (highlighted in yellow).

The highest age-gender group in the West Bank was 50–59-year females, while in Gaza it was 80+ males (highlighted in red).

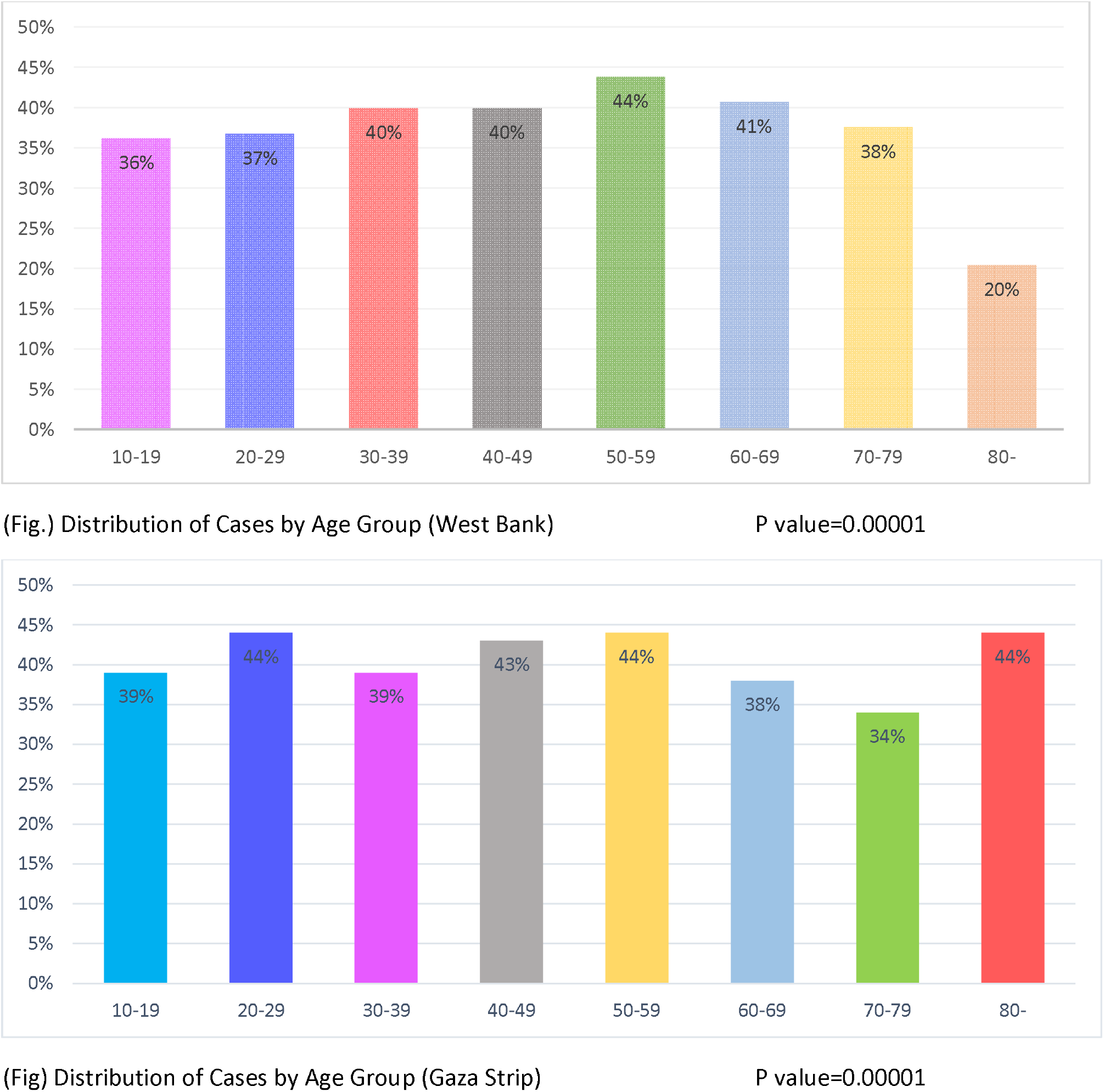

The two graphs above show that the difference between age groups, while statistically significant, does not show any massive disparities. The only interesting anomaly is that the 80+ group in the West Bank had a prevalence of less than half that of the 50-59 age group. This could be explained by policies implemented in the West Bank to protect the elderly or the adoption of specific individual behavior in the West Bank towards the elderly.

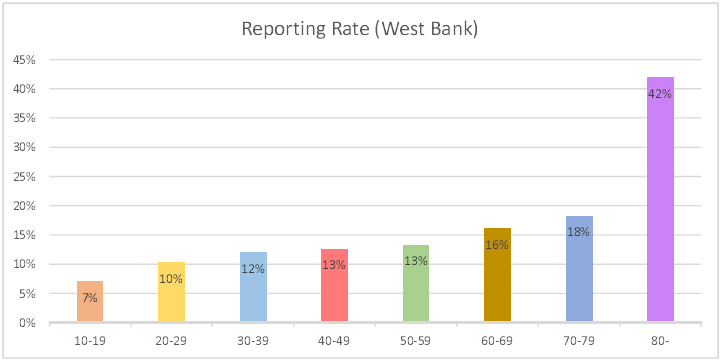

The Chart above shows the reporting rate for each age group. This is calculated by comparing the estimated number of infections (using the sero prevalence) to the number of cases reported during the same period of time. This graph shows that only 7% of infections were reported as cases for those between 10-19 years old, while 42% of infections were reported as cases for those over the age of 80.

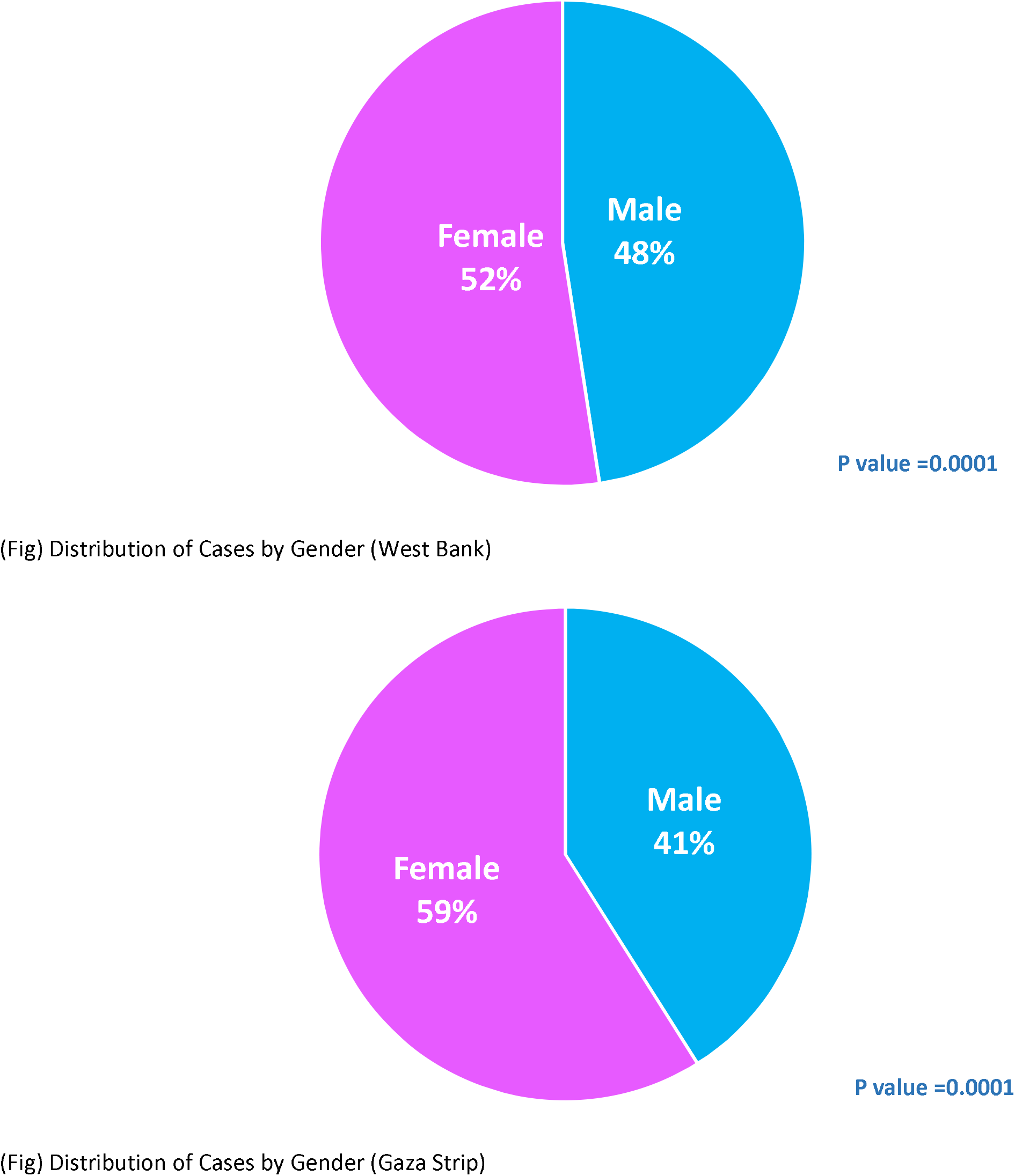

In both the West Bank and Gaza, seroprevalence in females was much higher than in males.

The area of residence had a significant effect as shown in the following table.

| Gender Distribution within Class (Adjusted for Sample) |  |  |  |
| --- | --- | --- | --- |
|  | Male | Female | Total |
| **West Bank** |  |  |  |
| **Camp** | 31% | 59% | **45%** |
| **Rural** | 30% | 43% | **36%** |
| **Urban** | 37% | 48% | **42%** |
| **Gaza** |  |  |  |
| **Camp** | 32% | 45% | **38%** |
| **Urban** | 40% | 43% | **41%** |

**P value = 0.0001**

Refugee camps in both the West Bank and Gaza had the highest prevalence. This is to be expected as camps have the
highest population density of any residential area.

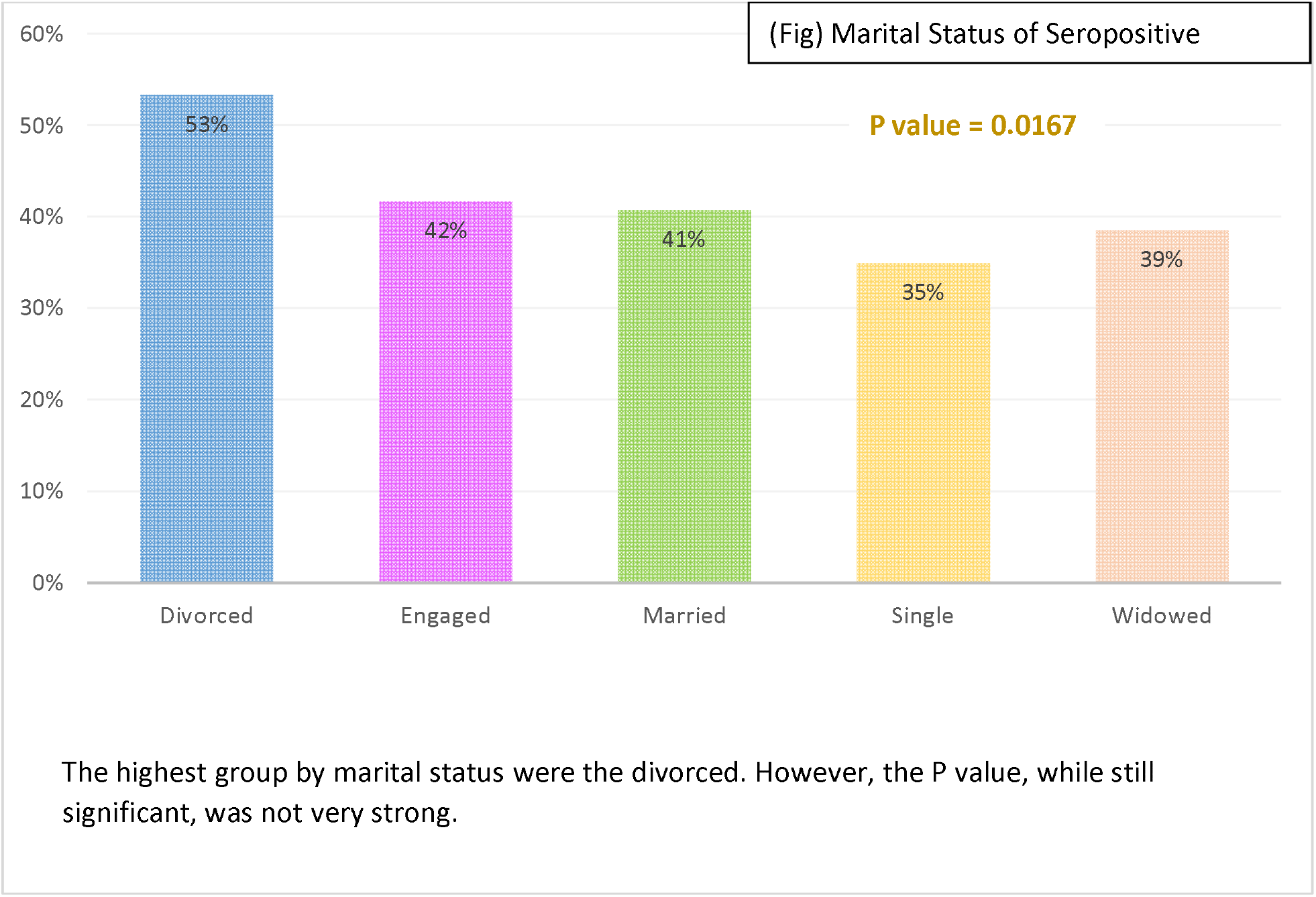

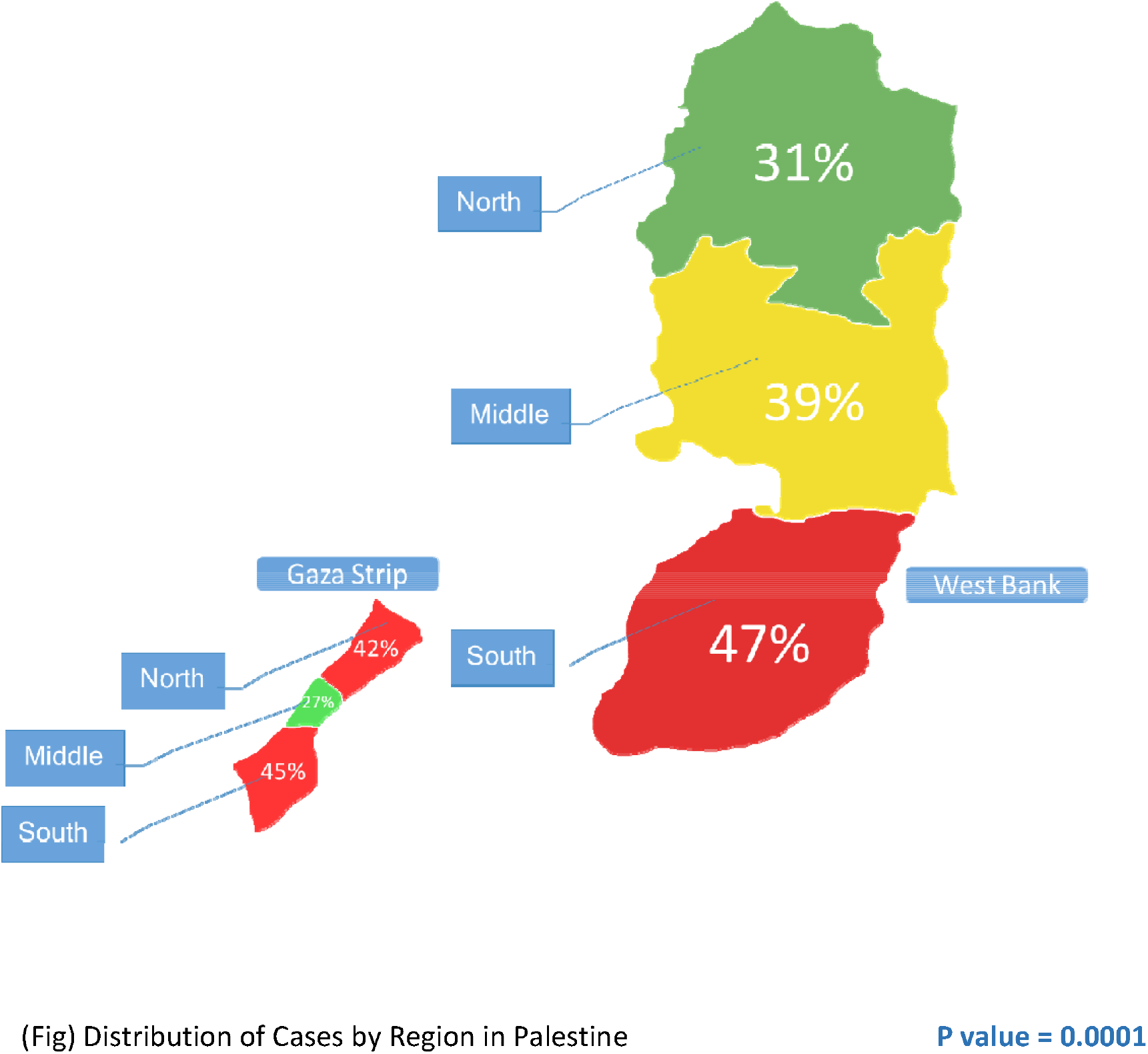

The different regions in the West Bank showed an interesting disparity in seroprevalence. There is a clear south to north pattern, with the south showing a prevalence 16% higher than the north.

Gaza’s regions indicated greater disparity with the southern region showing 45% prevalence, almost twice that of the middle region.

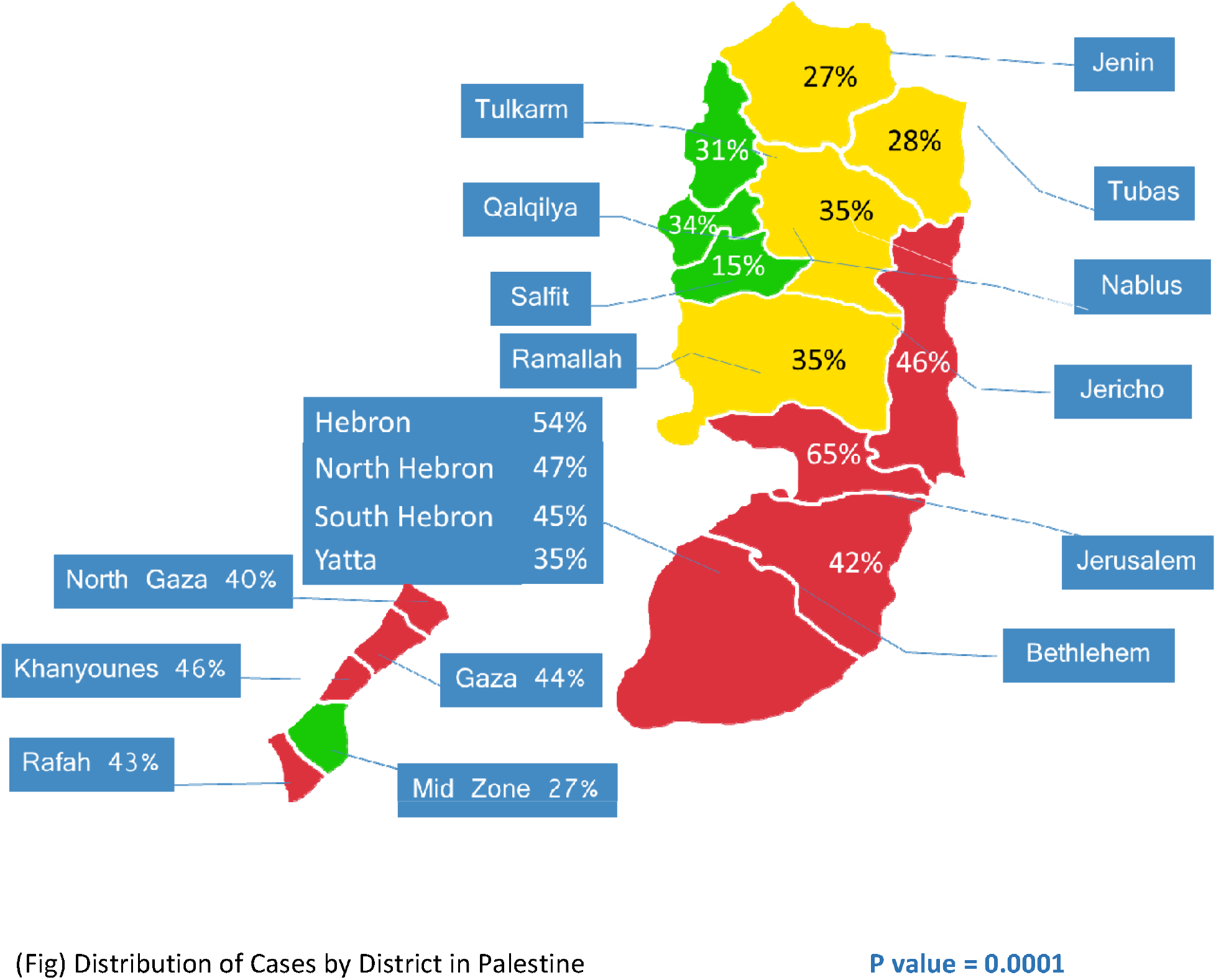

Prevalence in West Bank and Gaza districts showed a large variation in numbers. The Hebron district had the highest prevalence in Palestine at 54%, while Salfit had the lowest at 15%.

The mid-zone district in Gaza (or Deir Al Balah) had a low prevalence compared to the rest of the Gaza Strip. This can be explained by its lower population density of 3670/km^2^ compared to the Gaza district with 7092/km^2^.

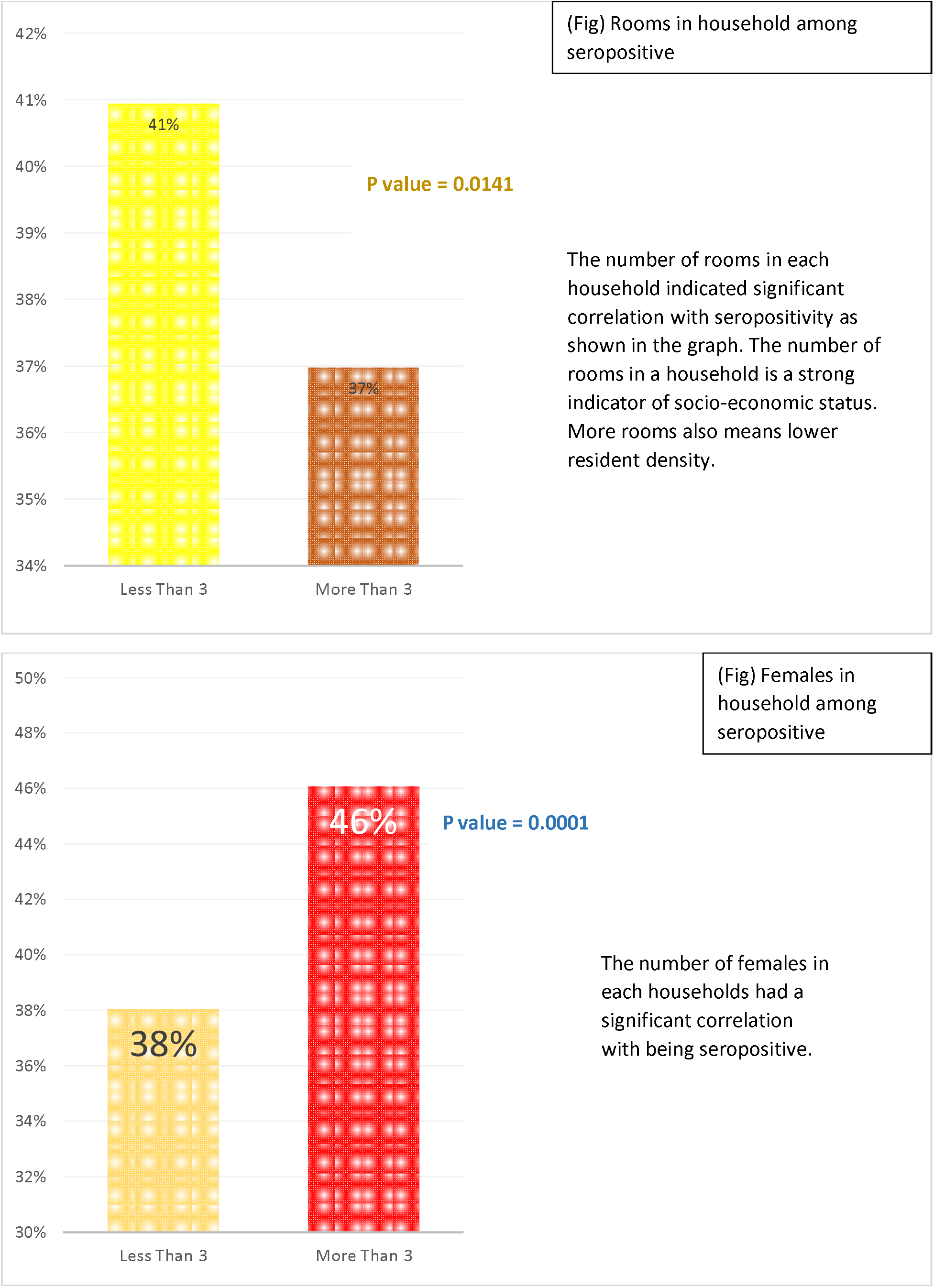

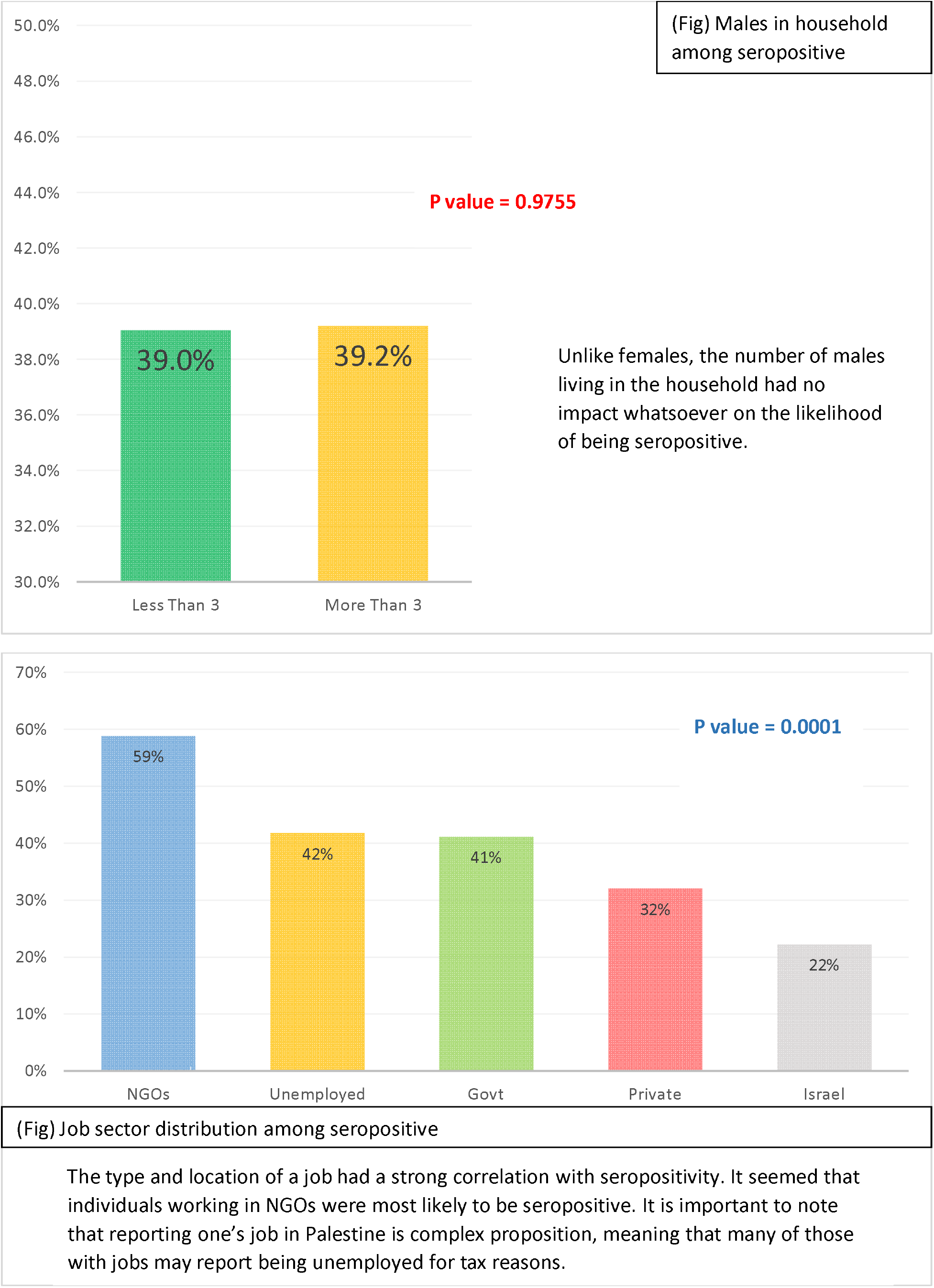

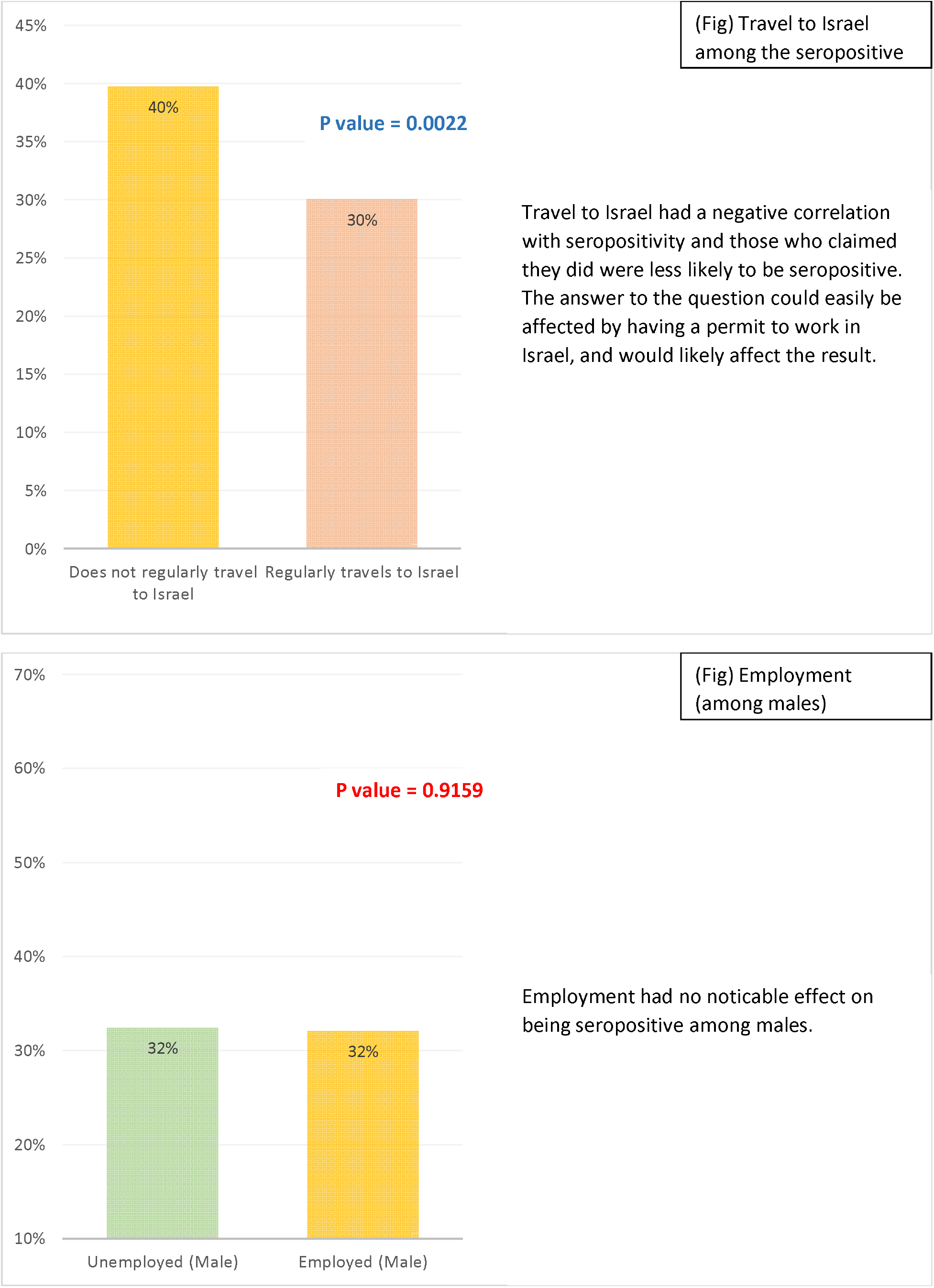

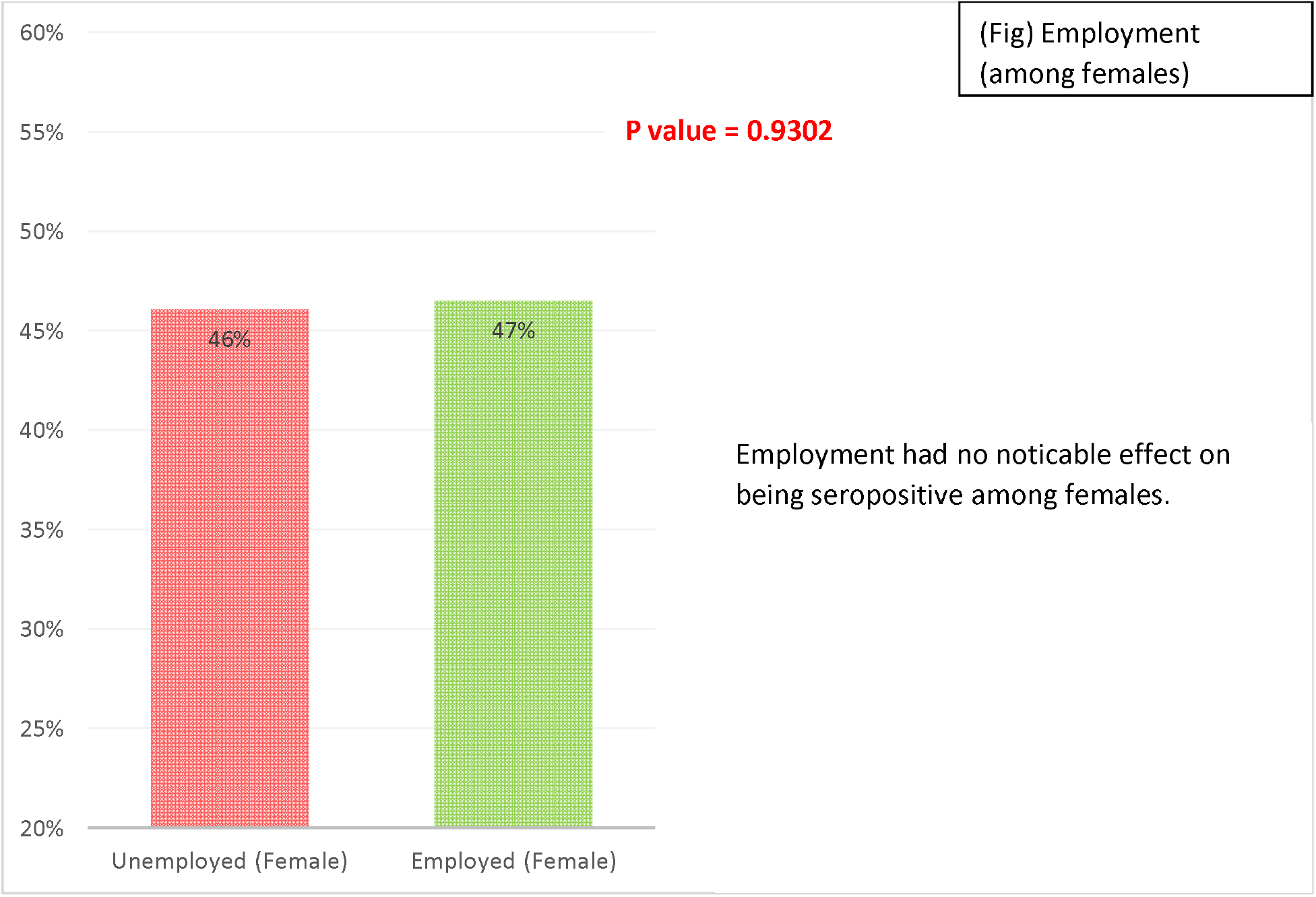

### Underlying Conditions and Risk Factors

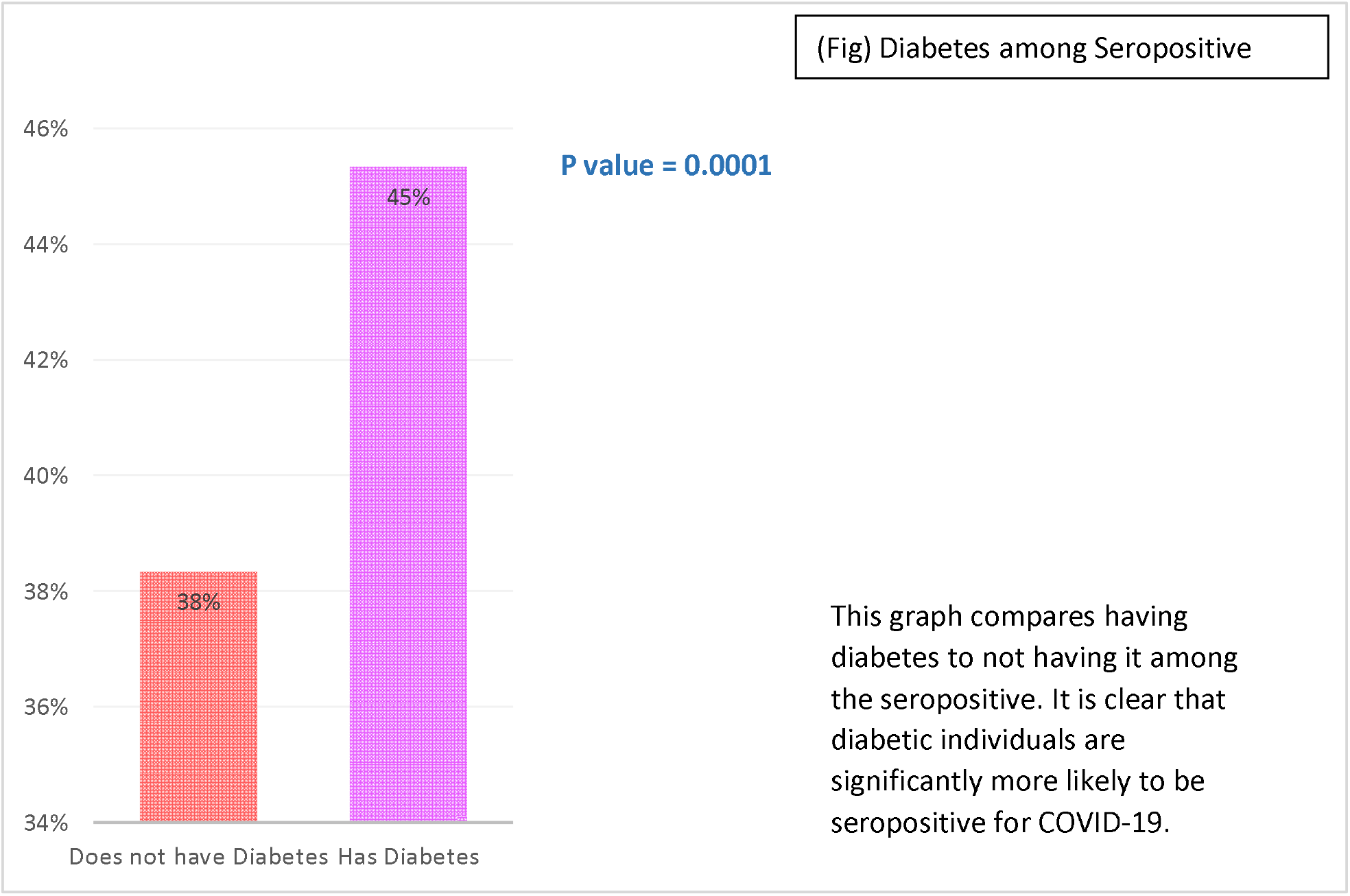

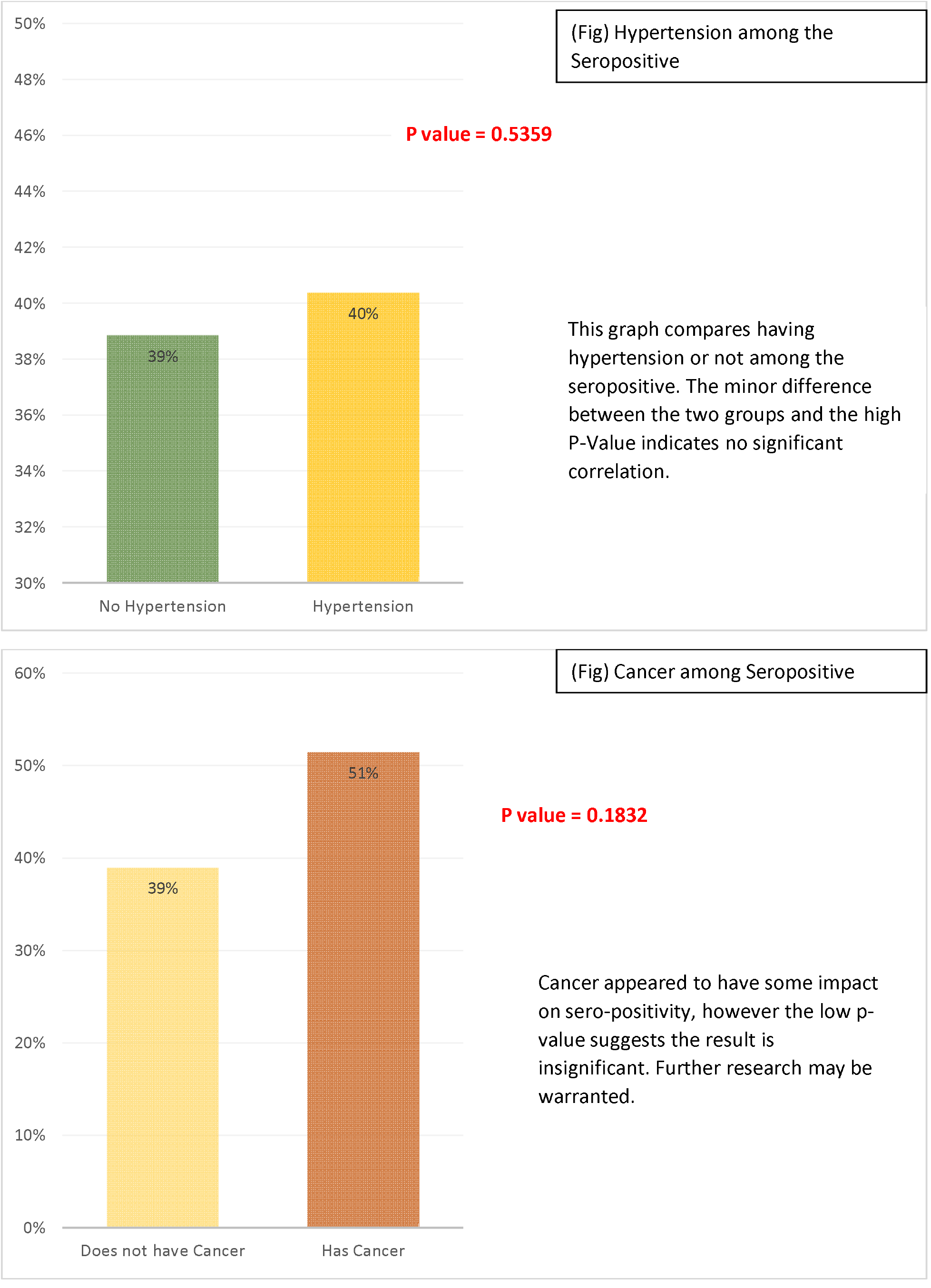

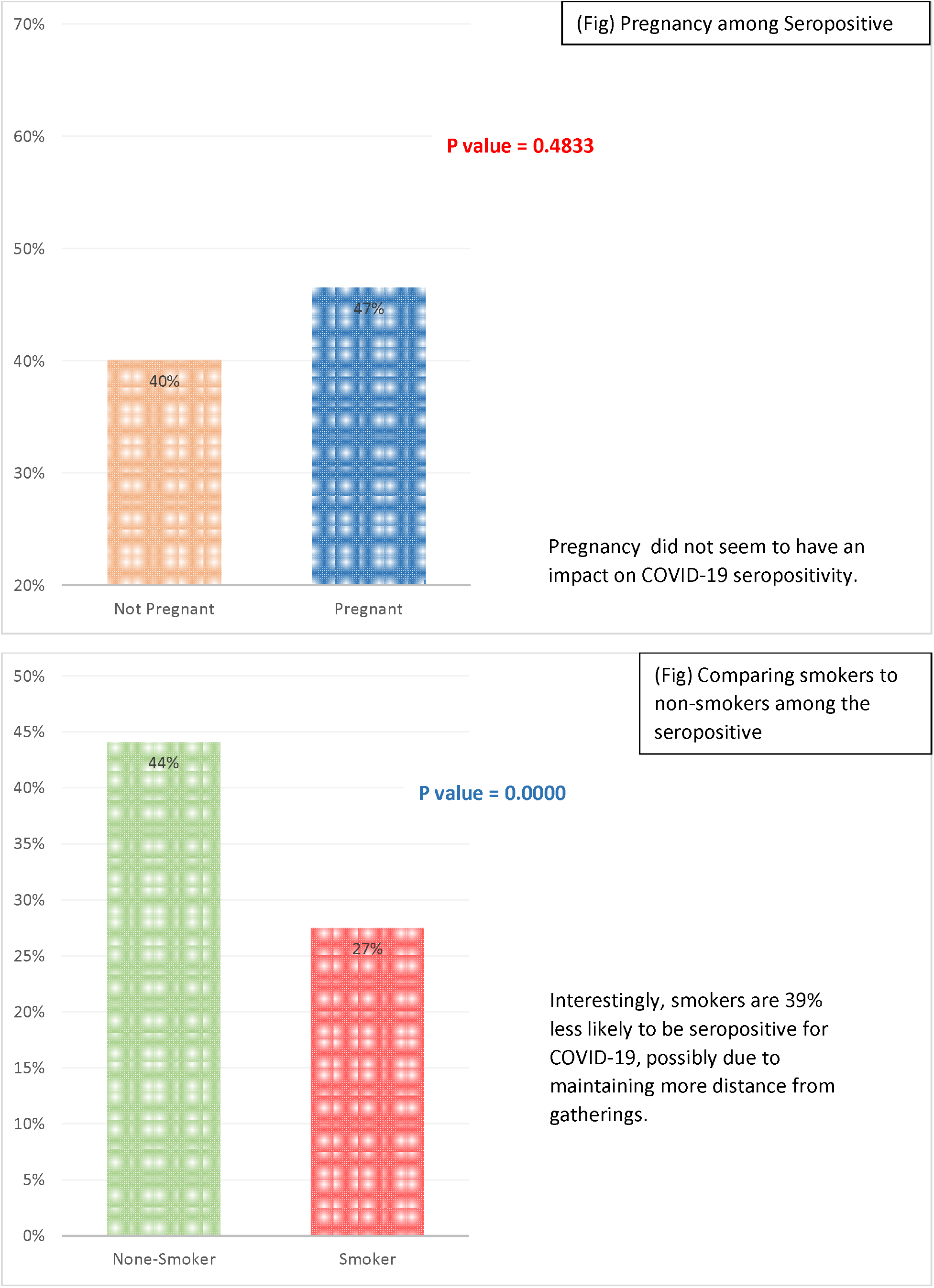

### Symptoms and detection

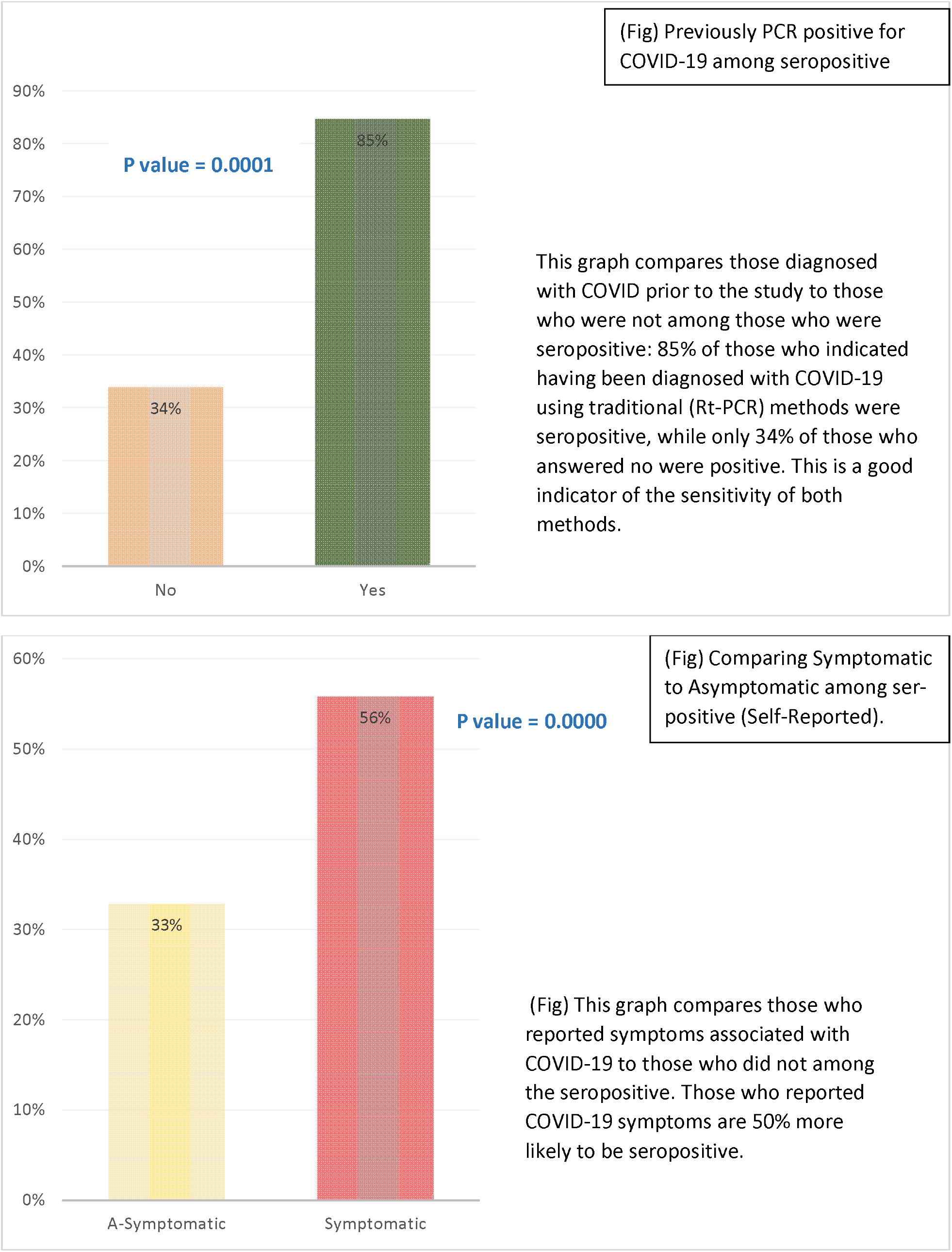

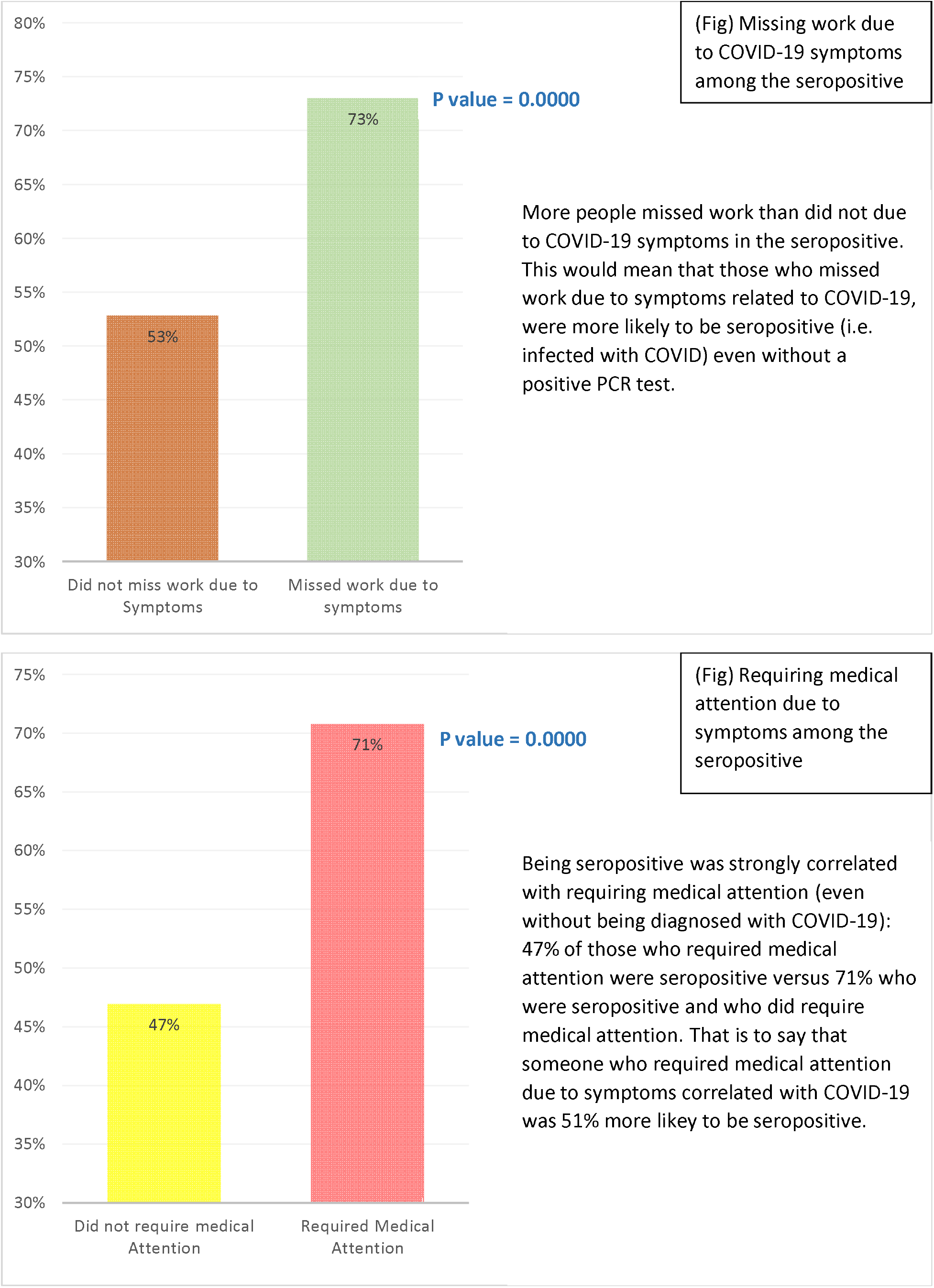

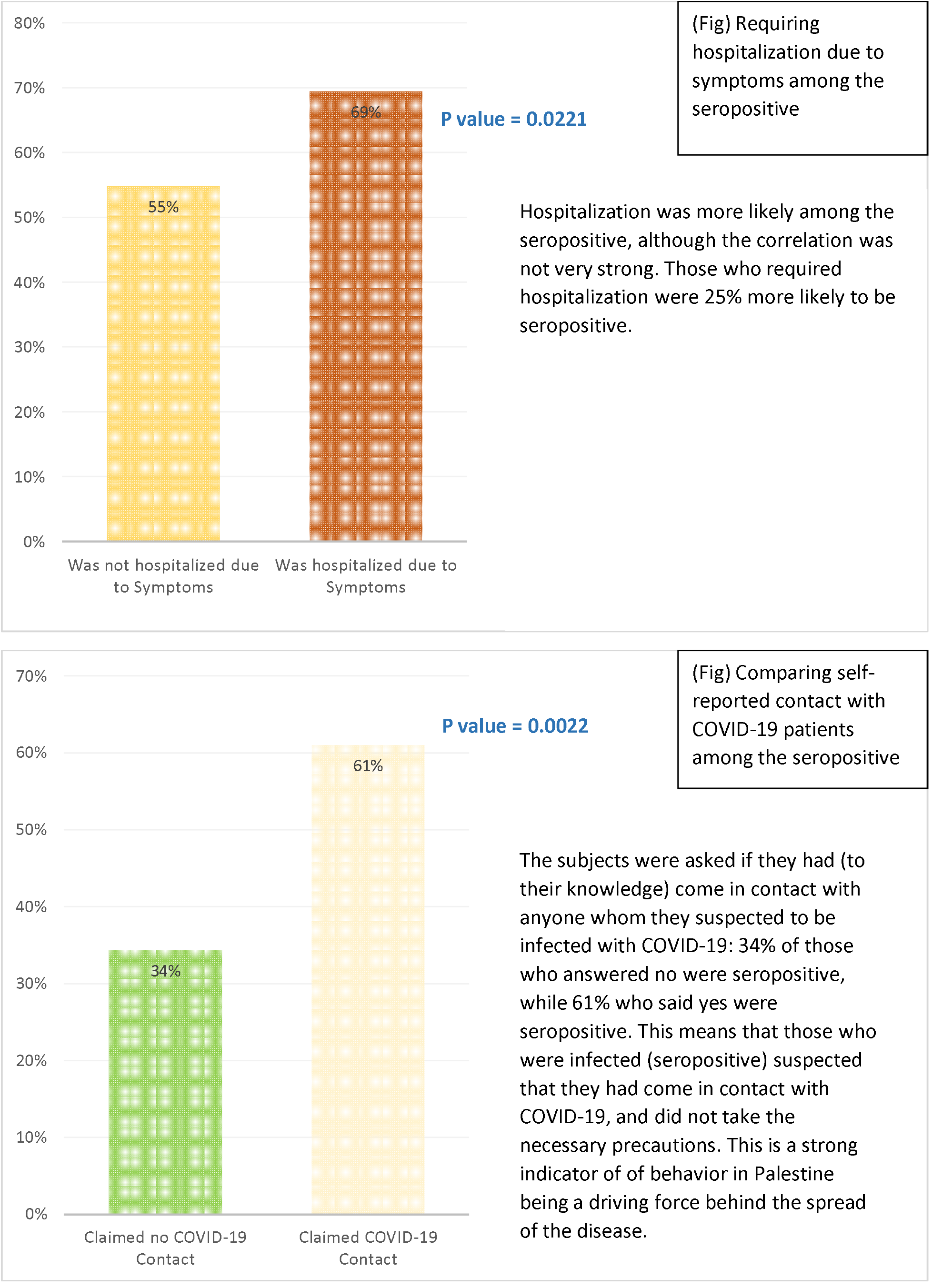

### Hospitalization Indicators

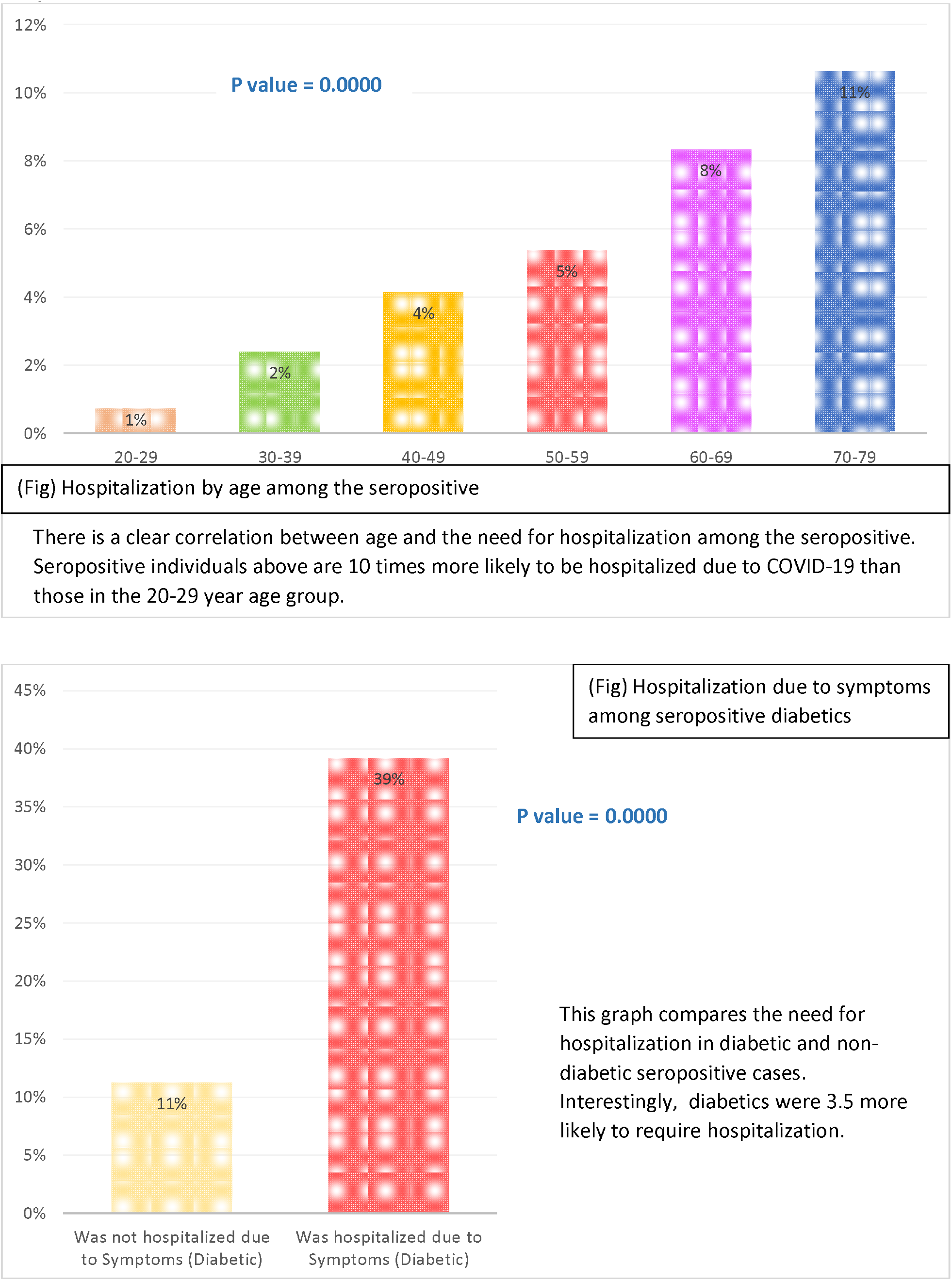

## Discussion

The results show clearly that a large percentage of the Palestinian population in the West Bank and Gaza had been infected with COVID-19 before the study was conducted. While rt-PCR tests indicated that 4% of the population had been infected by the end of 2020, sero-testing indicated that nearly 40% of the population had been detected within the same time frame, meaning that rt-PCR testing had detected only 10% of cases.

More females than males were infected across Palestine. Age was a strong factor in seroprevalence, with the 50–59-year age group being especially at risk compared to other groups. Interestingly, the 80+ age group in the West Bank was at much lower risk of being seropositive. It is possible that senior citizens adopted greater precautions in the West Bank such as social distancing and self-isolating, which reflects well on awareness campaigns. Such measures may not have been as easy to implement in Gaza due to the much higher population density of the region.

Reporting rates showed that the testing strategy heavily favors older citizens, which should come as no surprise as the testing strategy is test those who are symptomatic, and older people tend to be more symptomatic.

The type of residence was a major factor in seroprevalence. Refugee camps in the West Bank were at much higher risk than rural areas. Marital status was a factor as married or divorced individuals were at higher risk than single ones. Geographical factors were a major contributor, especially in the West Bank where southern districts were far more affected than northern ones. Southern districts have a higher population density and more close-knit communities that would increase the likelihood of infection.

As expected, socio-economic status in general seemed to affect the likelihood of infection. This can be seen in relation to correlations such as the number of rooms in a given household and commuting to Israel. The number of rooms in a given household can be an indicator of the socio-economic status of a family: more rooms indicate higher income; fewer rooms can indicate lower income and a higher density of residents in the household. In essence, those who answered yes to the question of traveling often to Israel were most likely workers with legal permits who tend to hold well-paid positions and other benefits that their “illegal” counterparts would not. Unfortunately, further research into the issue is difficult because answering this question truthfully could jeopardize the livelihoods of some individuals.

The presence of underlying health conditions seemed to have little effect on seroprevalence. However, diabetics appeared to have an increased risk of infection (1.2 times more likely). This is not to say that complications due to COVID infection are not affected by underlying conditions, but that the risk of becoming infected seems to be independent from underlying conditions. Diabetics are at more risk of infections in general because they are more likely to become hyperglycemic, which impairs the immune response. In addition, some diabetes-related health issues such as nerve damage and reduced blood flow to the extremities increase the body’s vulnerability to infection.^1^

Smoking appeared to have a negative effect on the likelihood of becoming infected and smoking reduced the chance of infection. This was rather unexpected as smoking is known to weaken the immune system^2^. It is possible that smokers maintain a greater distance from other people, meaning they could be better at social distancing.

Around 85% of subjects who had previously been diagnosed as being COVID-19 positive using rt-PCR were seropositive. They were 2.5 times more likely to be seropositive than those who had not been diagnosed. This did not seem to be affected by how long ago the detection occurred, as the graph below shows.

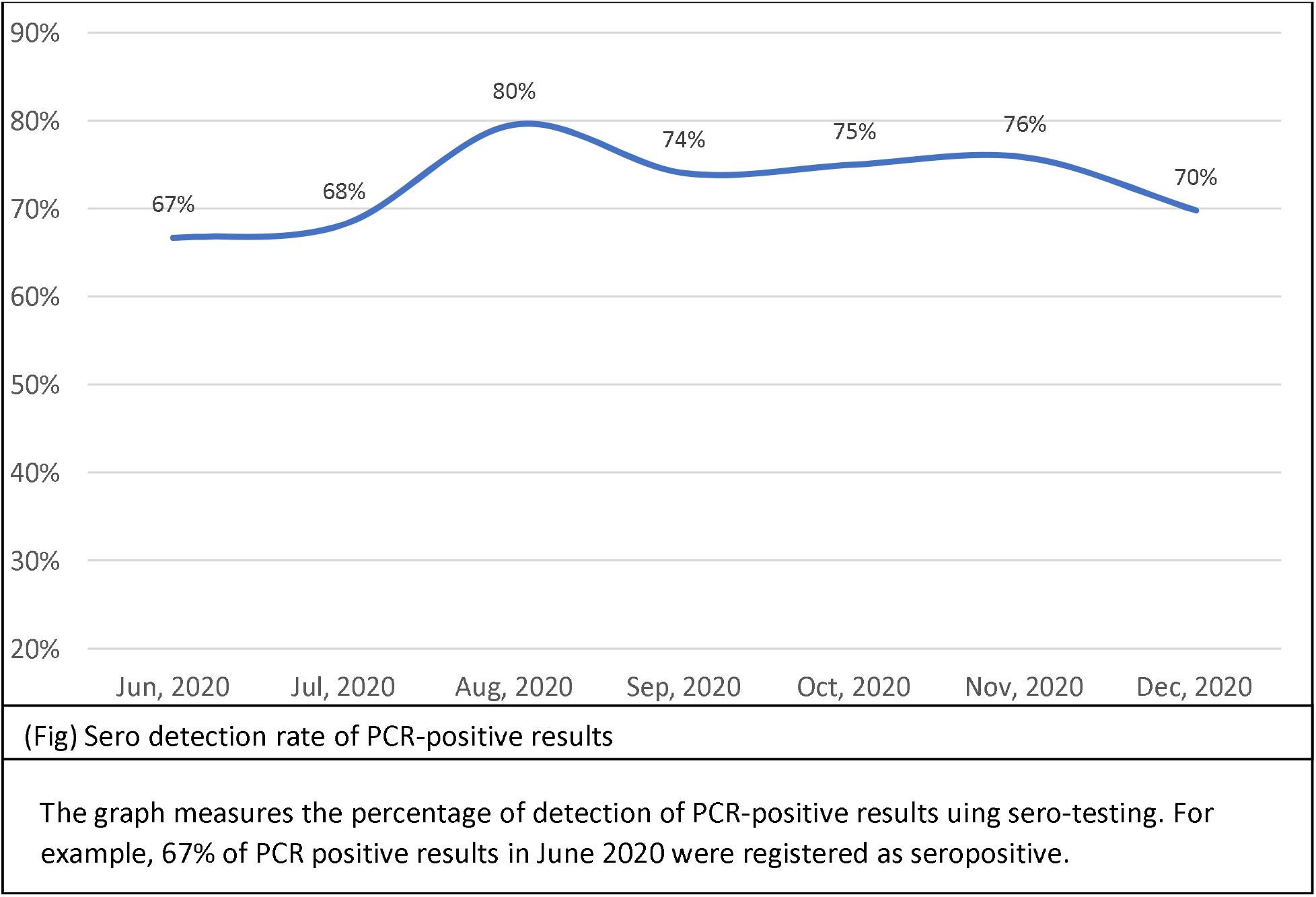

People who reported even one single symptom associated with COVID-19 were 1.7 times more likely to be seropositive. Those who missed work were 1.38 times more likely to be seropositive. Those who required medical attention were 1.51 more likely to be seropositive, and those who required hospitalization were 1.25 times more likely to be seropositive. These numbers indicate that being asymptomatic was more likely that previously thought, and that most undetected cases were actually mildly symptomatic, some of them severe enough to force the subject to miss work, require medical attention or even hospitalization.

Interestingly, people who reported suspecting that they had come in contact with COVID-positive individuals were 1.79 times more likely to be seropositive. This indicates that subjects were probably aware that they had been exposed.

Among those who were hospitalized, age was the key indicator. Those in the 70–79-year age group required hospital care 11 times more than those in the 20–29-year age group. Diabetics were 3.55 times more likely to require hospitalization when infected than non-diabetics, making diabetics the highest risk group.

The Sero Survey results allow us to calculate the IFR (Infection Fatality Rate). This is different from the CFR (Case Fatality Rate) which only accounts for actual cases detected at the time using rt-PCR. In contrast, the IFR is an estimate of the actual number of infections in the population, whether detected or not. The CFR in Palestine is about 1%, which is the global average for COVID-19, while the IFR in Palestine is 0.11%. This assumes a high level of detection of COVID-19 deaths.

The IFR in Palestine is close to that of other countries, as seen below.

|  |  |
| --- | --- |
| Mumbai, India | 0.09% |
| **Palestine** | **0.11%** |
| Qatar | 0.01% |
| Belgium | 1.09% |
| Iran | 0.08% |

### Recommendations

The Sero Survey revealed that Palestine has 10 times more cases than previously thought, meaning that the epidemic reached far more people than previously anticipated.

The patterns that emerge are substantial and could explain recent drops in previously endemic areas such as Hebron or increases in other areas. Certain groups are at more risk than others: females, diabetics, older people, and low-income families. Specific behaviors have serious repercussions on the likelihood of infection: travel, type of employment, awareness of contact, and suspicion of infection. This means that individuals can significantly reduce the likelihood of infection if they are made aware of these factors. Employers need to be very diligent in conveying to employees who suspect infection or have any symptoms that they should work from home and self-isolate. Even suspecting infection or having any symptom at all has been shown to increase the likelihood of being positive.

As this is the first study of its kind, it can be seen as a baseline for future studies. For example, it can be used to contrast a seroprevalence study conducted after a large-scale vaccination project has been concluded.

### Limitations

Several factors may have limited our ability to collect specimens. Certain communities were more resistant to testing than others due to the political realities in Palestine. Those same political factors also hindered our data collectors from travelling between different areas in the West Bank.

The geopolitical situation makes analysis of samples from Gaza and the West Bank in one single laboratory impossible. Each sample had to be analyzed in a lab in the same region where the sample had been collected. This could have a created a confounding effect when comparing Gaza with the West Bank.

## Data Availability

All data is included in the manuscript

## Acknowledgements

This study was made possible with the guidance and assistance of the World Health Organization’s East Mediterranean Regional Office, the Palestinian Ministry of Health, and with help from the Palestinian Central Bureau of Statistics.

## Footnotes

1 Berbudi A, Rahmadika N, Tjahjadi AI, Ruslami R. Type 2 Diabetes and its Impact on the Immune System. Curr Diabetes Rev. 2020;16(5):442-449. doi: 10.2174/1573399815666191024085838. PMID: 31657690; PMCID: PMC7475801.

2 Qiu F, Liang CL, Liu H, Zeng YQ, Hou S, Huang S, Lai X, Dai Z. Impacts of cigarette smoking on immune responsiveness: Up and down or upside down? Oncotarget. 2017 Jan 3;8(1):268-284. doi: 10.18632/oncotarget.13613. PMID: 27902485; PMCID: PMC5352117.

